# The relationship between diabetes, obesity and cardiovascular disease phenotypes: a UK Biobank cohort study

**DOI:** 10.1101/2023.01.03.23284157

**Authors:** Oliver I Brown, Michael Drozd, Hugo McGowan, Marilena Giannoudi, Marcella Conning-Rowland, John Gierula, Sam Straw, Stephen B Wheatcroft, Katherine Paradine, Lee D Roberts, Eylem Levelt, Ramzi Ajjan, Kathryn J Griffin, Marc A Bailey, Mark T Kearney, Richard M Cubbon

**Affiliations:** Leeds Institute for Cardiovascular and Metabolic Medicine, University of Leeds, Clarendon Way, Leeds, LS2 9JT, United Kingdom

**Author notes:** Author for correspondence, **Address for correspondence:** Dr Richard M Cubbon, LIGHT Laboratories 7.04, University of Leeds, Leeds, LS2 9JT, United Kingdom.

**Keywords:** Obesity, body mass index, diabetes mellitus, cardiometabolic, mortality, cardiovascular disease

## Abstract

**Introduction:** Obesity and diabetes mellitus (DM) frequently co-exist, yet their individual contributions to cardiovascular risk remain debated. We explored cardiovascular disease biomarkers, events and mortality in a large cohort stratified by body mass index (BMI) and DM.

**Methods:** 451,355 UK Biobank participants were stratified by ethnicity-specific BMI categories (normal, overweight, obese) and DM status. We examined cardiovascular biomarkers including: carotid intima-media thickness (CIMT); arterial stiffness; left ventricular ejection fraction (LVEF) and cardiac contractility index (CCI). Poisson regression models estimated adjusted incidence rate ratios (IRR) for myocardial infarction, ischaemic stroke and cardiovascular death, with normal weight non-DM as comparator.

**Results:** 5% of participants had DM (10% normal weight, 34% overweight and 55% obese; versus 34%, 43% and 23%, respectively, in non-DM). In the non-DM group, overweight/obesity was associated with higher CIMT, arterial stiffness and CCI, and lower LVEF (p<0.05); these relationships were diminished in the DM group. Within BMI classes, DM was associated with adverse cardiovascular phenotype (p<0.05), particularly in the normal weight group. After 5,323,190 person-years follow-up, incident myocardial infarction, ischaemic stroke and cardiovascular mortality all rose across increasing BMI categories in the DM and non-DM groups (p<0.05). However, normal weight DM had higher cardiovascular mortality than obese non-DM (IRR 2.81 [95% confidence interval: 2.24-3.54] vs IRR 1.84 [1.70-1.98]).

**Conclusions:** Obesity and DM are additively associated with adverse cardiovascular biomarkers and mortality risk. Whilst adiposity metrics are more strongly correlated with cardiovascular biomarkers than diabetes-oriented metrics, both correlate weakly, suggesting other factors underpin the high cardiovascular risk of normal-weight diabetes.

## Introduction

Diabetes mellitus (DM) and obesity are major causes of cardiovascular morbidity and mortality worldwide. [1] Obesity is a complex condition of increased subcutaneous and visceral adiposity, often associated with adipose dysfunction and insulin resistance, which increases the risk of DM and cardiovascular disease. [2–4] Most people with DM are overweight or obese, except for the minority with autoimmune or genetic forms of diabetes, and for each unit increase in body mass index (BMI), the likelihood of DM increases exponentially. [5–7] DM and obesity are associated with increased vascular stiffness and accelerated atherosclerosis, processes which lead to premature cardiovascular disease and death. [8–10]

Large population studies attempting to discern the independent cardiovascular risk conferred by DM suggest that ‘adjusting’ for BMI does not substantially diminish the association between DM and cardiovascular mortality. [11] However, the relation between BMI and cardiovascular disease is potentially complex, with BMI above or below the normal range being associated with higher risk of cardiovascular (and all-cause) mortality. [12,13] Such data may reflect residual confounding factors and suggest cautious interpretation of epidemiological data in isolation. Cardiovascular imaging studies, whilst much smaller, offer an alternative approach to characterise overt and subclinical cardiovascular disease in people with DM and/or obesity. These support the notion that DM in the context of ‘normal’ BMI is still associated with important cardiovascular abnormalities.[14,15]

The literature defining the complex relationship between DM, BMI and cardiovascular disease lacks data leveraging multimodality cardiovascular imaging and hard outcomes within a single cohort of sufficient size to study people with normal BMI and DM. To address this, we used the UK Biobank (UKB) cohort study. We hypothesised that DM with normal BMI would be associated with a cardiovascular phenotype and event rate comparable to obesity without DM.

## Methods

### Study population

UKB is a prospective observational cohort study of 502,462 participants aged between 37 to 73 years, recruited from 22 assessment centres across the United Kingdom (UK) between 2006 and 2010. It is an open access resource developed using UK Government and biomedical research charity funding which collected genotypic, phenotypic and linked health care record data. Full details of the study design and conduct are available from the UKB website (https://www.ukbiobank.ac.uk). The UKB received ethical approval from the NHS Research Ethics Service (11/NW/0382); we conducted this analysis under application number 59585. All participants provided written informed consent.

### Definitions of diabetes, body mass index and study covariates

Baseline sociodemographic characteristics, comorbidities and medication were recorded by participants completing a touchscreen and nurse-led interview, as previously described. [16] Data extracted from a face-to-face nurse-led interview was used to categorise participants comorbidities and medication use at recruitment. DM was classified as any self-reported diagnosis of “diabetes” (1220); “type 1 diabetes mellitus” (1222); “type 2 diabetes mellitus” (1223); “diabetic eye disease” (1276); “diabetic neuropathy/ulcers” (1468) and “diabetic nephropathy” (1607). Duration of diabetes was defined as the time from self-reported diagnosis to the date of study recruitment.

BMI category was adjusted for ethnicity in accordance with World Health Organisation (WHO) ethnicity-specific threshold recommendations: normal: BMI ≥18.5kg/m^2^ to <25 kg/m^2^ or ≥18.5kg/m^2^ to <23 kg/m^2^ if South Asian ethnicity; overweight: ≥25kg/m^2^ to <30 kg/m^2^ or ≥23kg/m^2^ to <27.5 kg/m^2^ if South Asian ethnicity; obese: ≥30kg/m^2^ or ≥27.5kg/m^2^ if South Asian ethnicity. [17] Participants with below normal BMI were excluded from this analysis (n=2316) due to an insufficient sample size to study people with DM. Definitions of other comorbidities at recruitment has previously been described and is shown in appendix 1. [18] We excluded participants with missing data relating to BMI (n=10,135), loss to follow-up or withdrawal consent (n=1298), or confounding factors: smoking status (n=2949); ethnicity (n=2777); socioeconomic status (n=624) and systolic blood pressure (n=34,439, SBP).

### Assessment of cardiometabolic phenotype

From 2014, all surviving participants were invited by email and then post, to take part in multimodality imaging assessment. This included cardiac magnetic resonance imaging (cMRI), carotid artery ultrasound, photoplethysmography derived arterial stiffness index (PASI), abdominal MRI, anthropometric measurements of body composition and measurements of serum lipids and biochemistry. Responding participants were screened for eligibility for inclusion based on safety and tolerability (i.e. claustrophobia and inability to breath hold) criteria. All participants with metal implants in their body (including MRI compatible devices) were excluded for safety and concerns about degrading image quality. [19] Full details of the protocol for each method of assessment have been previously described. [20–25]

cMRI was conducted with standardised protocols in a subpopulation of 35,972 participants. Image acquisition was performed during a 20-min protocol performed using a 1.5T scanner (MAGNETOM Aera, Syngo Platform VD13A, Siemens Healthcare, Erlangen, Germany). [20] Detailed assessment of cardiac structure and function was performed including bright blood anatomic assessment (sagittal, coronal, and axial); left and right ventricular cine images, myocardial tagging; native T1 mapping and aortic flow. However, at the time of this analysis only data related to left ventricular ejection fraction (LVEF), left ventricular end diastolic volume (LVEDV), left ventricular end systolic volume (LVESV), left ventricular stroke volume (LVSV) and cardiac output was available for our entire subpopulation. We indexed measurements according to body surface area (BSA) and computed cardiac contractility index (CCI), which is derived from SBP divided by indexed LVESV and is a validated measure of global myocardial contractility which is inversely associated with mortality among patients with heart failure.[26–28]

Carotid artery ultrasound was performed in 41,442 participants using a CardioHealth Station (Panasonic Healthcare Corporation of North America, Newark, USA) with a 9 MHz linear array transducer.[21] Carotid intima-media thickness (CIMT) was performed at two angles for each carotid, giving a total of four CIMT measurements from which mean CIMT was calculated. PASI was measured in 162,029 participants using the PulseTrace PCA2 (CareFusion; San Diego, CA). PASI (in m/s) was calculated by dividing waveform standing height by the time between forward and the reflected waves of the pulse waveform detected from an infrared sensor on the index finger over a 15 second period. [22] SBP, DBP and heart rate were measured in the entire study population at recruitment using the Omron Digital blood pressure monitor (Omron, Kyoto, Japan). Two measurements were taken in each participant from which the mean was calculated.

Abdominal MRI was performed using a Siemens Aera 1.5T scanner (Syngo MR D13) (Siemens, Erlangen, Germany). The imaging protocol covered a 1.1m region of the study participants from neck-to-knee region, including some organs outside the abdominal cavity. A single 3D volume for each participant using an automated fat-water swap detection and correction procedure was calculated. [23] Data relating to visceral adipose tissue volume (VAT), subcutaneous adipose tissue volume (SAT) and total thigh fat-free muscle volume was available for 9,407 participants at the time of analysis. Total abdominal adipose tissue index (TAATI) was defined as VAT volume + abdominal SAT volume / BSA. Abdominal fat ratio (AFR) was defined as VAT volume + SAT volume / VAT volume + abdominal SAT volume + total thigh fat-free muscle volume.[29] Anthropometric data was available on 450,798 participants during the baseline visit. Weight and bioimpedance were measured using the Tanita BC418ma bioimpedance device (Tanita, Tokyo, Japan), from which percentages of body fat and lean mass were estimated. [24] Non-fasted venous blood sampling was performed and were processed at a central laboratory.

### Definition of mortality, nonfatal myocardial infarction, and ischaemic stroke

UKB mortality outcomes are obtained from the official UK national death registry from National Health Service (NHS) digital for participants in England and Wales, and from the NHS central register for participants in Scotland. We censored outcome data to the 23^rd^ of March 2021. Cardiovascular death was defined according to the International Classification of Diseases, Tenth Revision (ICD-10) as previously described.[30] In brief, this included all cardiovascular ICD-10 codes from I00-I99 excluding codes relating to infection death. Secondary outcomes included incident non-fatal myocardial infarction (MI) or incident non-fatal ischaemic stroke which were defined using an algorithm developed by UKB which linked hospital admissions data with related ICD codes and patient reported outcomes. [31,32] We only included outcome data ascertained from hospital admission data (excluding self-reported outcomes) and where non-fatal MI or non-fatal ischaemic stroke was the primary diagnosis.

### Statistical analysis

All analyses were performed using Stata/MP. All statistical tests were 2-sided and statistical significance was defined as p<0.05. Missing data were not imputed. Continuous data are presented as median with 25^th^-75^th^ centile. Categorical data are presented as number (%). Normality of distribution was checked using skewness and kurtosis tests and all continuous variables were found to be non-normally distributed. Differences between BMI categories within the diabetes or the non-diabetes group were assessed using Kruskal-Wallis H tests or Chi^2^ test for continuous and categorical variables, respectively. Differences between the diabetes and non-diabetes groups within each BMI category were assessed using the Mann-Whitney U tests. Where appropriate, some analyses were repeated after stratification by sex. We used correlation matrices of Poisson model’s coefficients to assess correlations between covariates; no correlation coefficients higher than 0·3 or lower than –0·3 were observed.

Unadjusted and adjusted incident rate ratios (IRR) and their 95% confidence intervals (CI) were estimated for all-cause mortality, cardiovascular mortality, non-fatal MI, and ischaemic stroke using Poisson regression models; exposure time was modelled, but time-varying covariates were not used. A dummy variable was used to compare outcomes of DM-BMI groups with reference to the normal BMI non-DM group. Where indicated, models were adjusted for age, sex, ethnicity, and smoking status. Crude mortality rates were calculated per 1000 participant-years of follow-up for all-cause mortality, cardiovascular mortality, non-fatal MI, and ischaemic stroke by ethnicity-adjusted BMI category for the total population and then stratified by diabetes status. Kaplan-Meier (KM) curves for clinical outcomes were used to illustrate unadjusted event rates among participants grouped by diabetes status and ethnicity-adjusted BMI category. Where specified, we separately modelled BMI as a continuous variable using restricted cubic splines with four knots for all clinical outcomes, as this provided the best fit as assessed by minimising Akaike and Bayesian criteria (models including categorical, linear, or cubic splines with three, four, and five knots and first-degree and second-degree fractional polynomials were compared). Models were constructed independently for participants with and without diabetes. The reference knot was set at the median BMI of the whole cohort and spline curves were truncated at the 1^st^ and 99th centile.

## Results

We included 451,355 participants of whom 22,451 (4.9%) had DM at recruitment. Among participants with DM 2,290 (10.1%) were normal weight, 7,732 (34.4%) overweight and 12,429 (55.3%) obese. In participants without DM, 143,557 (33.5%) were normal weight, 185,758 (43.3%) overweight and 99,589 (23.2%) obese. Participants with DM were older, more often male, less physically active, less often of non-white ethnicity and more socio-economically deprived (**Table 1**). There was greater prevalence of all studied cardiometabolic comorbidities at baseline among participants with DM, and the prevalence of cardiometabolic comorbidities (except for peripheral vascular disease) increased with BMI, irrespective of DM status (table 1). Antihypertensive, statin, aspirin and diuretic use was higher among participants with DM at enrolment and usage increased with increasing BMI category regardless of DM status. Most participants with DM were taking metformin and approximately a fifth were receiving insulin (**Table S1**). DM medication use (except for insulin) rose with increasing BMI.

**Table 1.** Population demographic characteristics and comorbidities at study recruitment. Participants stratified by diabetes status and then by ethnicity adjusted BMI category. Normal: BMI ≥18.5kg/m^2^ to <25 kg/m^2^ *or* ≥18.5kg/m^2^ to <23 kg/m^2^ if south Asian ethnicity; Overweight: ≥25kg/m^2^ to <30 kg/m^2^ *or* ≥23kg/m^2^ to <27.5 kg/m^2^ if south Asian ethnicity; Obese: ≥30kg/m^2^ *or* ≥27.5kg/m^2^ if south Asian ethnicity. Continuous data presented as median with 25^th^ and 75^th^ centile. Categorical data presented as n (%). * represents Kruskal Wallis p value or chi^2^ test <0.05 between BMI categories for continuous variables and categorical variables respectively within diabetes or non-diabetes groups. ^†^ represents p value <0.05 between each BMI category in diabetes participants and their respective BMI category in non-diabetes participants from Mann Whitney U test and chi^2^ test for categorical variables. Abbreviations: metabolic equivalent (MET), body mass index (BMI). Where fewer than 50 participants are within any group, UK Biobank requires that the specific number of participants is not listed to reduce the risk of de-anonymisation.

|  |  | No Diabetes |  |  | Diabetes |  |  |
| --- | --- | --- | --- | --- | --- | --- | --- |
|  | Missing | Normal<br>(N=143 557) | Overweight<br>(N=185 758) | Obese<br>(N=99 589) | Normal<br>(N=2290) | Overweight<br>(N=7732) | Obese<br>(N=12 429) |
| *Demographic characteristics* |  |  |  |  |  |  |  |
| Age, years | 0 | 56 (49-62)*† | 58 (50-63)*† | 58 (50-63)*† | 61 (55-66)*† | 62 (57-66)*† | 61 (51-65)*† |
| Male sex, n (%) | 0 | 49 099 (34.2)*† | 96 862 (52.1)*† | 45 938 (46.1)*† | 1326 (57.9)*† | 5366 (69.4)*† | 7317 (58.9)† |
| Ethnicity, n (%) | 0 |  |  |  |  |  |  |
| White | - | 138 120 (96.2)*† | 176 483 (95.1)*† | 92 500 (92.9)*† | 2006 (87.6)*† | 6569 (85.0)*† | 10 958 (88.2)*† |
| Mixed | - | 964 (0.7)*† | 1011 (0.5)*† | 606 (0.6)*† | <50 (0.7)*† | 56 (0.7)*† | 81 (0.7)*† |
| Asian | - | 1147 (0.8)*† | 3448 (1.9)*† | 2895 (2.9)*† | 98 (4.3)*† | 600 (7.8)*† | 775 (6.2)*† |
| Black | - | 1320 (0.9)*† | 2804 (1.5)*† | 2566 (2.6)*† | 88 (3.8)*† | 301 (3.9)*† | 422 (3.4)*† |
| Chinese | - | 857 (0.6)*† | 400 (0.2)*† | 69 (0.1)*† | <50 (1.2)*† | <50 (0.5)*† | <50 (0.1)*† |
| Other | - | 1149 (0.8)*† | 1613 (0.9)*† | 953 (1.0)*† | 55 (2.4)*† | 170 (2.2)*† | 179 (1.4)*† |
| Townsend Deprivation Index, n (%) | 0 |  |  |  |  |  |  |
| Q1 | - | 31 140 (21.7)*† | 39 054 (21.0)*† | 16 960 (17.0)*† | 398 (17.4)*† | 1254 (16.2)*† | 1648 (13.3)*† |
| Q2 | - | 29 964 (20.9)*† | 38 622 (20.8)*† | 18 241 (18.3)*† | 423 (18.5)*† | 1368 (17.7)*† | 1864 (15.0)*† |
| Q3 | - | 28 771 (20.0)*† | 38 063 (20.5)*† | 19 275 (19.4)*† | 428 (18.7)*† | 1488 (19.2)*† | 2125 (17.1)*† |
| Q4 | - | 28 053 (19.5)*† | 36 481 (19.6)*† | 21 012 (21.1)*† | 468 (20.4)*† | 1590 (20.6)*† | 2632 (21.2)*† |
| Q5 | - | 25 629 (17.9)*† | 33 538 (18.1)*† | 24 101 (24.2)*† | 573 (25.0)*† | 2032 (26.3)*† | 4160 (33.5)*† |
| Smoking, n (%) | 0 |  |  |  |  |  |  |
| Never | - | 84 895 (59.1)*† | 100 138 (53.9)*† | 52 108 (52.3)*† | 1187 (51.8)*† | 3476 (45.0)*† | 5541 (44.6)*† |
| Former | - | 42 604 (29.7)*† | 66 762 (35.9)*† | 37 896 (38.1)*† | 780 (34.1)*† | 3388 (43.8)*† | 5656 (45.5)*† |
| Current | - | 16 058 (11.2)*† | 18 858 (10.2)*† | 9585 (9.6)*† | 323 (14.1)*† | 868 (11.2)*† | 1232 (9.9)*† |
| Summed MET activity per week, minutes | 86 677 | 2009 (975-3813)*† | 1813 (847-3625)*† | 1428 (594-3110)*† | 1769 (822-3550)*† | 1536 (677-3200)*† | 1184 (450-2754)*† |
| *Cardiometabolic diseases* |  |  |  |  |  |  |  |
| Hypertension, n (%) | 0 | 20 184 (14.1)*† | 46 104 (24.8)*† | 38 399 (38.6)*† | 1023 (44.7)*† | 4588 (59.3)*† | 8947 (72.0)*† |
| Atrial fibrillation/flutter, n (%) | 0 | 678 (0.5)*† | 1317 (0.7)*† | 869 (0.9)*† | <50 (1.0)*† | 71 (0.9)*† | 172 (1.4)*† |
| Peripheral vascular disease n (%) | 0 | 524 (0.4)† | 643 (0.4)† | 396 (0.4)† | <50 (1.0)† | 93 (1.2)† | 116 (0.9)† |
| Stroke or transient ischaemic attack, n (%) | 0 | 1562 (1.1)*† | 2831 (1.5)*† | 2140 (2.2)*† | 73 (3.2)*† | 340 (4.4)*† | 585 (4.7)*† |
| Heart failure, n (%) | 0 | 98 (0.1)*† | 180 (0.1)*† | 164 (0.2)*† | <50 (0.3)† | <50 (0.3)† | 56 (0.5)† |
| Ischaemic heart disease, n (%) | 0 | 2961 (2.1)*† | 7644 (4.1)*† | 5965 (6.0)*† | 243 (10.6)*† | 1208 (15.6)*† | 2196 (17.7)*† |
| Chronic cardiac syndrome, n (%) | 0 | 3065 (2.1)*† | 7824 (4.2)*† | 6108 (6.1)*† | 252 (11.0)*† | 1224 (15.8)*† | 2234 (18.0)*† |
| Aortic aneurysmal disease, n (%) | 0 | 74 (0.1)* | 142 (0.1)*† | 106 (0.1)*† | <50 (0)* | <50 (0.3)*† | <50 (0.2)*† |
| *Non-cardiometabolic diseases* |  |  |  |  |  |  |  |
| Chronic liver disease, n (%) | 0 | 223 (0.2)* | 345 (0.2)*† | 204 (0.2)*† | <50 (0.3) | <50 (0.3)† | 53 (0.4)† |
| Chronic respiratory disease, n (%) | 0 | 16 708 (11.6)*† | 22 857 (12.3)* | 15 436 (15.5)*† | 298 (13.0)*† | 989 (12.8)* | 2206 (17.8)*† |
| Chronic renal disease, n (%) | 0 | 288 (0.2)† | 405 (0.2)† | 237 (0.4)† | <50 (1.1)† | 61 (0.8)† | 116 (0.9)† |
| Neurological disease, n (%) | 0 | 1829 (1.3)* | 2303 (1.2)* | 1381 (1.4)* | <50 (1.6) | 89 (1.2) | 154 (1.2) |
| Psychiatric disease, n (%) | 0 | 7356 (5.1)*† | 10 255 (5.5)* | 7816 (7.9)*† | 150 (6.6)*† | 449 (5.8)* | 1097 (8.8)*† |
| Rheumatological disease, n (%) | 0 | 2810 (2.0)*† | 3935 (2.1)*† | 2621 (2.6)*† | 71 (0.3)*† | 192 (2.5)*† | 391 (3.2)*† |

### Phenotypic measures of metabolic disease

Participants with DM had elevated BMI, waist to hip ratio (WHR), body fat percentage and reduced whole body impedance compared to those without diabetes within any given BMI category (**Table 2**). Increasing BMI category was associated with significantly higher serum triglycerides, low-density lipoprotein cholesterol (LDL) and lower high-density lipoprotein cholesterol (HDL) irrespective of DM status. Total cholesterol, LDL, HDL and triglycerides were lower in participants with diabetes compared to those without within any given BMI category (**Table 2**). Participants with DM had higher serum creatinine, serum cystatin C, urinary microalbumin and serum alanine aminotransferase compared to those without DM, which rose with increasing BMI category. Higher BMI categories were also associated with increased serum c-reactive protein (CRP) and HbA1c regardless of DM status (**Table 2**). Within any given BMI category, TAATI and AFR were elevated in participants with DM, compared to those without; both measures increased in higher BMI groups (**Table 2**). Given the sexual dimorphism in body composition, we also performed stratified analyses which show similar patterns in relation to DM and BMI category in both sexes (**Table S2**). Collectively, these data illustrate important differences in metabolic parameters associated with increasing BMI, irrespective of DM status; however, many of these are also abnormal in people with DM and ‘normal’ BMI versus those without DM.

**Table 2.** Metabolic phenotypes of study participants. Participants stratified by diabetes status and then by ethnicity adjusted BMI category. Normal: BMI ≥18.5kg/m^2^ to <25 kg/m^2^ *or* ≥18.5kg/m^2^ to <23 kg/m^2^ if south Asian ethnicity; Overweight: ≥25kg/m^2^ to <30 kg/m^2^ *or* ≥23kg/m^2^ to <27.5 kg/m^2^ if south Asian ethnicity; Obese: ≥30kg/m^2^ *or* ≥27.5kg/m^2^ if south Asian ethnicity. Continuous data presented as median with 25th and 75th centile. Categorical data presented as n (%). * represents Kruskal Wallis p value between BMI categories for continuous variables within diabetes or non-diabetes groups. ^†^ represents p value <0.05 between each BMI category in diabetes participants and their respective BMI category in non-diabetes participants from independent t-tests for continuous variables and chi^2^ test for categorical variables. Abbreviations: glycated haemoglobin (HbA1c); high density lipoprotein (HDL); insulin-like growth factor 1 (IGF-1); low density lipoprotein (LDL).

|  |  | No Diabetes |  |  | Diabetes |  |  |
| --- | --- | --- | --- | --- | --- | --- | --- |
|  | Missing | Normal<br>(N=143 557) | Overweight<br>(N=185 758) | Obese<br>(N=99 589) | Normal<br>(N=2290) | Overweight<br>(N=7732) | Obese<br>(N=12 429) |
| Diabetes metrics |  |  |  |  |  |  |  |
| Diabetes duration, years | 159 | - | - | - | 8 (3-19)* | 6 (2-11)* | 5 (2-10)* |
| Age of diabetes diagnosis, years | 159 | - | - | - | 52 (37-59)* | 55 (46-60)* | 53 (46-59)* |
| Body composition |  |  |  |  |  |  |  |
| BMI | 0 | 23.1 (21.8-24.1)*† | 27.1 (26.0-28.4)*† | 32.6 (31.1-35.1)*† | 23.4 (22.3-24.3)*† | 27.7 (26.4-28.8)*† | 34.0 (31.7-37.5)*† |
| Waist circumference, cm | 63 | 78 (72-84)*† | 91 (85-97)*† | 104 (97-110)*† | 84 (77-89)*† | 96 (90-101)*† | 110 (103-118)*† |
| Hip circumference, cm | 56 | 96 (93-99)*† | 103 (100-106)*† | 112 (108-118)*† | 96 (92-99)*† | 102 (99-105)*† | 113 (108-120)*† |
| Waist to hip ratio | 88 | 0.81 (0.76-0.87)*† | 0.89 (0.82-0.94)*† | 0.92 (0.85-0.98)*† | 0.88 (0.82-0.93)*† | 0.94 (0.89-0.98)*† | 0.97 (0.91-1.03)*† |
| Body fat percentage | 219 | 28 (22-33)*† | 31 (25-38)*† | 40 (31-45)*† | 24 (20-30)*† | 28 (25-35)*† | 37 (32-44)*† |
| Whole body fat mass, kg | 557 | 17.2 (14.1-20.4)*† | 23.9 (20.4-27.5)*† | 34.8 (29.9-40.6)*† | 16.2 (13.2-19.5)*† | 23.3 (20.1-26.7)*† | 36.0 (30.5-43.4)*† |
| Whole body impedance, ohms | 37 | 656 (597-710)*† | 585 (532-648)*† | 539 (488-597)*† | 617 (565-675)*† | 559 (517-612)*† | 507 (464-561)*† |
| Lipids |  |  |  |  |  |  |  |
| Serum Apolipoprotein A, g/L | 65 270 | 1.6 (1.4-1.8)*† | 1.5 (1.3-1.7)*† | 1.4 (1.3-1.6)*† | 1.5 (1.3-1.7)*† | 1.4 (1.2-1.6)*† | 1.3 (1.2-1.5)*† |
| Serum Apolipoprotein B, g/L | 29 354 | 1.0 (0.8-1.1)*† | 1.0 (0.9-1.2)*† | 1.1 (0.9-1.2)*† | 0.8 (0.7-0.9)*† | 0.8 (0.7-1.0)*† | 0.8 (0.7-1.0)*† |
| Serum total cholesterol, mmol/L | 27 198 | 5.7 (5.0-6.4)*† | 5.8 (5.0-6.5)*† | 5.7 (4.9-6.5)*† | 4.4 (3.8-5.1)*† | 4.3 (3.8-5.1)*† | 4.3 (3.8-5.0)*† |
| Serum HDL cholesterol, mmol/L | 63 179 | 1.6 (1.3-1.9)*† | 1.4 (1.2-1.6)*† | 1.3 (1.1-1.5)*† | 1.4 (1.1-1.7)*† | 1.2 (1.0-1.4)*† | 1.1 (0.9-1.3)*† |
| Serum LDL cholesterol, mmol/L | 28 008 | 3.4 (2.9-4.1)*† | 3.6 (3.1-4.2)*† | 3.6 (3.0-4.2)*† | 2.5 (2.1-3.0)*† | 2.5 (2.1-3.1)*† | 2.6 (2.2-3.1)*† |
| Serum lipoprotein A, nmol/L | 112 118 | 20.6 (9.5-60.3)* | 21.4 (9.7-61.6)* | 21.5 (9.5-64.3)*† | 19.8 (8.8-60.1)* | 21.4 (9.0-67.2)* | 18.7 (8.3-62.2)*† |
| Serum triglycerides, mmol/L | 27 550 | 1.2 (0.9-1.6)* | 1.6 (1.1-2.2)*† | 1.9 (1.3-2.6)*† | 1.2 (0.8-1.7)* | 1.7 (1.2-2.5)*† | 2.0 (1.5-2.8)*† |
| Biochemistry |  |  |  |  |  |  |  |
| Creatinine, μmol/L | 27 425 | 67 (59-76)*† | 73 (63-83)*† | 72 (63-82)*† | 70 (60-81)*† | 73 (64-85)*† | 72 (62-84)*† |
| Serum cystatin C, mg/L | 27 240 | 0.84 (0.77-0.92)*† | 0.89 (0.81-0.98)*† | 0.94 (0.86-1.04)*† | 0.87 (0.78-0.99)*† | 0.92 (0.83-1.04)*† | 0.97 (0.87-1.11)*† |
| Urinary microalbumin, mg/L | 314 521 | 10.7 (8.2-16.8)*† | 10.9 (8.2-17.6)*† | 12.1 (8.7-21.1)*† | 14.4 (9.2-29.7)*† | 14.9 (9.8-32.2)*† | 17.6 (10.3-42.2)*† |
| Alanine aminotransferase, U/L | 27 210 | 17 (14-22)*† | 21 (16-28)*† | 24 (18-33)*† | 21 (16-27)*† | 24 (18-32)*† | 26 (19-37)*† |
| C-reactive protein, mg/L | 28 117 | 0.8 (0.4-1.5)*† | 1.3 (0.7-2.5)* | 2.5 (1.4-4.7)*† | 0.9 (0.4-1.9)*† | 1.3 (0.7-2.6)* | 2.5 (1.2-4.9)*† |
| Diabetes related biomarkers |  |  |  |  |  |  |  |
| Glucose, mmol/L | 63 478 | 4.9 (4.5-5.2)*† | 4.9 (4.6-5.3)*† | 5.0 (4.7-5.4)*† | 6.4 (5.2-9.5)*† | 6.4 (5.3-8.6)*† | 6.6 (5.4-9.0)*† |
| HbA1c, mmol/mol | 29 788 | 34 (32-37)*† | 35 (33-37)*† | 36 (34-39)*† | 50 (43-59)*† | 50 (43-58)*† | 51 (44-60)*† |
| IGF-1, nmol/L | 29 518 | 21.7 (18.1-25.2)*† | 21.7 (18.1-25.2)*† | 20.1 (16.3-23.8)*† | 21.0 (17.1-25.2)*† | 20.9 (16.7-24.8)*† | 18.5 (14.4-22.8)*† |
| Abdominal MRI |  |  |  |  |  |  |  |
| Abdominal fat ratio, fraction | 442 119 | 0.44 (0.36-0.51)* | 0.51 (0.44-0.58)* | 0.60 (0.53-0.66)*† | 0.46 (0.45-0.57)* | 0.50 (0.45-0.57)* | 0.61 (0.56-0.67)*† |
| Total abdominal adipose tissue index, L/m<sup>2</sup> | 441 948 | 2.5 (1.9-3.2)* | 3.8 (3.1-4.6)* | 5.7 (4.7-6.8)*† | 2.6 (1.8-3.5)* | 3.8 (3.2-4.7)* | 6.1 (5.2-7.0)*† |

### Phenotypic measures of cardiovascular disease

Resting heart rate, SBP and DBP increased across rising BMI categories. Participants with DM also had higher resting heart rate and SBP but lower DBP than participants without DM within each BMI category (**Table 3**). Participants with DM had higher CIMT and PASI than those without BM within each BMI category; both measures increased with rising BMI in the non-DM group, but only PASI (not CIMT) increased with BMI in the DM group (**Table 3**). Among those who were overweight or obese, LVEF was lower in participants with DM compared to those without. Furthermore, LVEF declined with rising BMI category, irrespective of DM status (**Table 3**). Among those without DM, rising BMI was associated with lower LVSV, LVEDV, cardiac index; however, CCI increased with rising BMI. In participants with DM, rising BMI was only significantly associated with lower LVSV and higher CCI. Within any given BMI category, participants with DM had evidence of elevated CCI. Given the potential sexual dimorphism in these measures, we also performed stratified analyses which show similar patterns in relation to DM and BMI category in both sexes (**Table S3**). Collectively, these data illustrate an adverse cardiovascular phenotype associated with rising BMI in people without DM, although this relationship is less clear in people with DM who exhibit marked abnormalities even in the ‘normal’ BMI group. Pairwise correlation analysis demonstrated that duration of DM weakly correlated with CIMT and LVEF, but not PASI or CCI (**Figure 1**); no correlations were noted with HbA1c in the DM group, but in people without DM, HbA1c correlated with PASI, CIMT, LVEF and CCI. The strongest correlates of cardiovascular parameters were markers of adiposity; this was apparent in the DM and non-DM groups, although these remained modest with Pearson’s correlation coefficients of no more than 0.25.

**Table 3.**
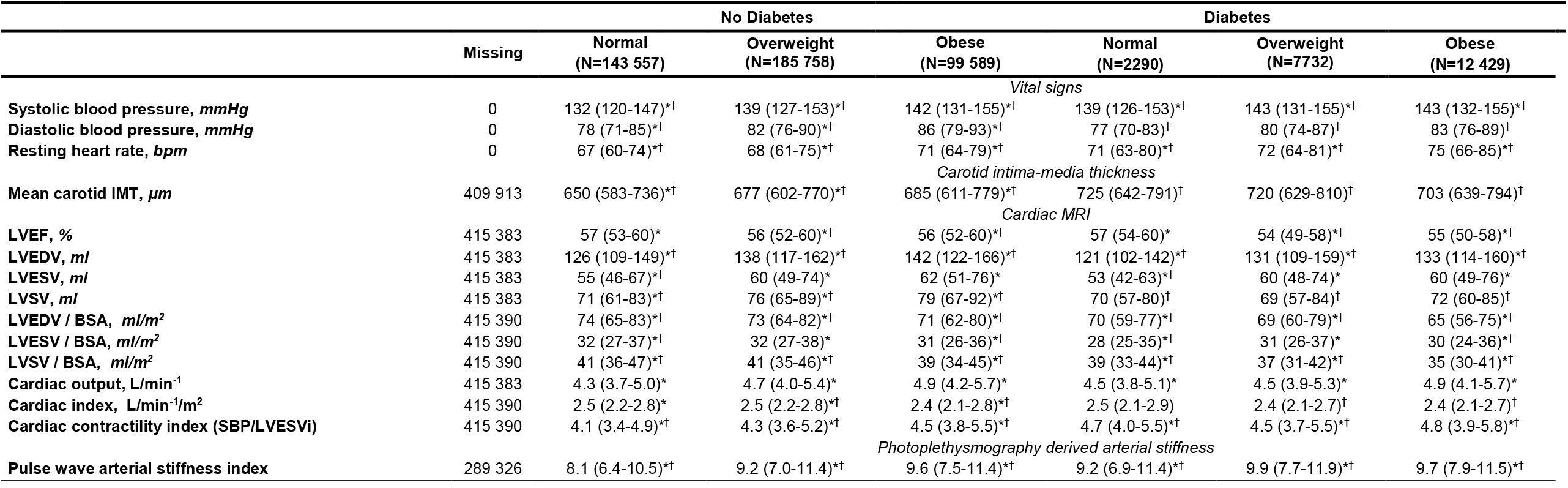
Phenotypic measurements of cardiovascular disease. Participants are stratified by diabetes status and then by ethnicity adjusted BMI category. Normal: BMI ≥18.5kg/m^2^ to <25 kg/m^2^ *or* ≥18.5kg/m^2^ to <23 kg/m^2^ if south Asian ethnicity; Overweight: ≥25kg/m^2^ to <30 kg/m^2^ *or* ≥23kg/m^2^ to <27.5 kg/m^2^ if south Asian ethnicity; Obese: ≥30kg/m^2^ *or* ≥27.5kg/m^2^ if south Asian ethnicity. Continuous data presented as median with 25^th^ and 75^th^ centile. Categorical data presented as n (%). * represents Kruskal Wallis p value or chi^2^ test <0.05 between BMI categories for continuous variables and categorical variables respectively within diabetes or non-diabetes groups. ^†^ represents p value <0.05 between each BMI category in diabetes participants and their respective BMI category in non-diabetes participants from Mann Whitney U tests for continuous variables and chi^2^ test for categorical variables. Abbreviations: body mass index (BMI) body surface area (BSA), intima media thickness (IMT), left ventricular ejection fraction (LVEF), left ventricular end-diastolic volume (LVEDV), left ventricular end-systolic volume (LVESV), left ventricular end-systolic volume indexed to body surface area (LVESVi), left ventricular stroke volume (LVSV), systolic blood pressure (SBP).

**Figure 1.**
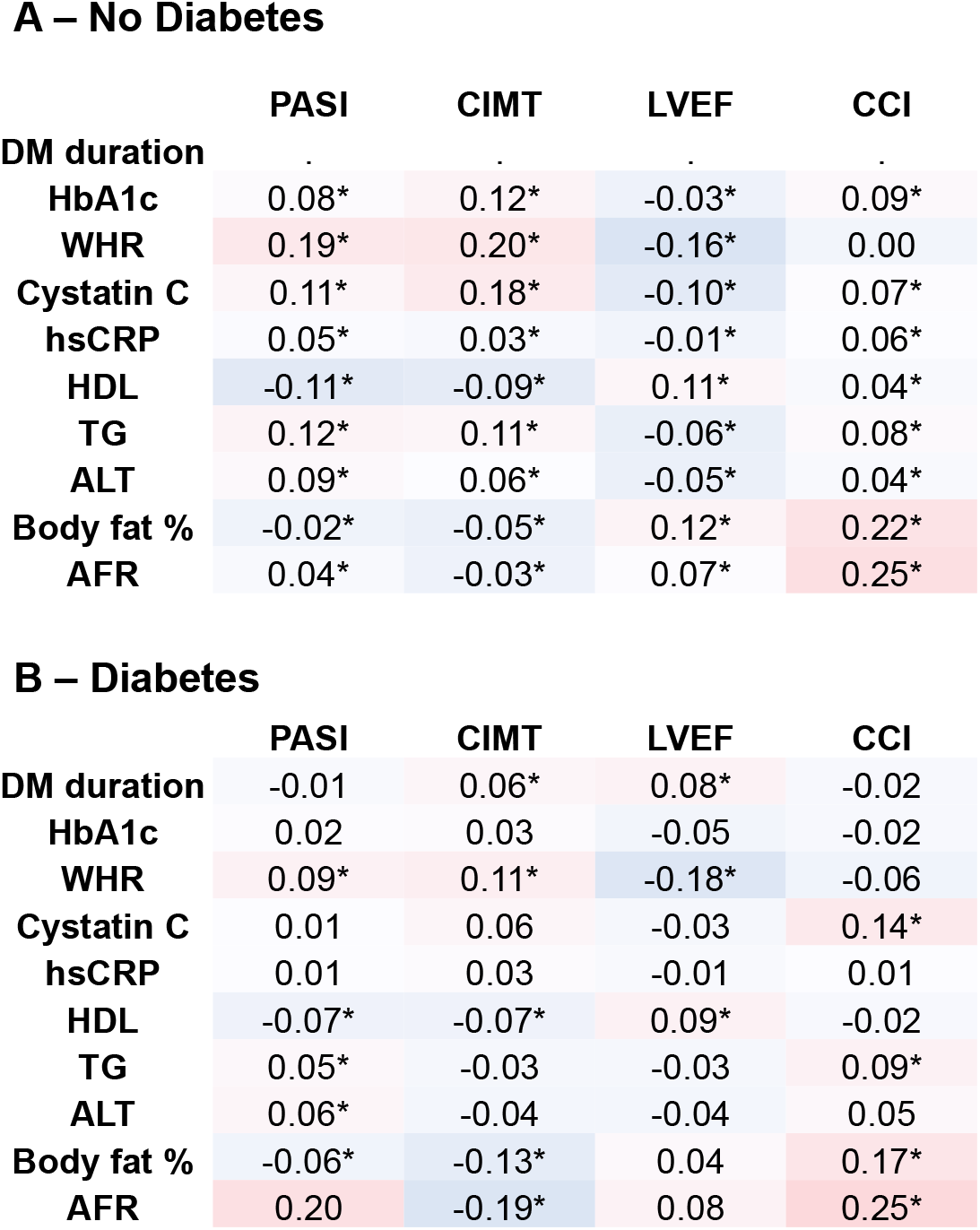
Correlation of cardiovascular imaging phenotypes with biomarkers of metabolic phenotype stratified by diabetes status. Numbers represent Pearson’s correlation coefficient with * indicating p<0.05. AFR is defined as visceral adipose tissue (VAT) + abdominal subcutaneous adipose tissue volume (SAT) / VAT volume + abdominal SAT volume + total thigh fat-free muscle volume. CCI is defined as systolic blood pressure / left ventricular end-systolic volume indexed to body surface area. Abbreviations: abdominal fat ratio (AFR); alanine aminotransferase (ALT); body surface area (BSA); cardiac contractility index (CCI); carotid intima media thickness (CIMT); diabetes mellitus (DM); glycated haemoglobin (HbA1c); high density lipoprotein (HDL); high sensitivity c-reactive protein (hsCRP); left ventricular ejection fraction (LVEF); pulse wave arterial stiffness index (PASI); subcutaneous adipose tissue (SAT); triglycerides (TG); visceral adipose tissue (VAT); waist to hip ratio (WHR).

### All-cause mortality, cardiovascular mortality and morbidity

During 5,323,190 person-years of follow-up (median 12.0 years), 29,931 participants died (6.6%) of whom 5,831 (1.3%) died from cardiovascular causes. A total of 7,179 (1.6%) participants had non-fatal MI and 3,469 (0.8%) had non-fatal ischaemic stroke during follow-up. Kaplan-Meier curves illustrating all-cause and cardiovascular mortality during follow-up are shown in **Figure 2**. Broadly, these demonstrate modestly rising mortality across BMI categories with much greater mortality in groups with DM. Indeed, absolute unadjusted rates of all-cause mortality, cardiovascular death, non-fatal MI and non-fatal ischaemic stroke climbed modestly with increasing BMI category, with much greater mortality in participants with DM (**Table S4**). Unadjusted and adjusted IRRs for all-cause and cardiovascular mortality are shown in **Table 4**. Among participants without DM, the risk of cardiovascular death was higher among overweight (adjusted IRR 1.09, CI 1.02-1.18) and obese participants (adjusted IRR 1.70, CI: 1.58-1.84) compared to those with normal BMI. However, people with DM and normal BMI experienced a near 2.5-fold (adjusted IRR 2.45, CI: 1.94-3.10) higher risk of cardiovascular death with further increases in the overweight DM (adjusted IRR 2.90, CI: 2.56-3.28) and obese DM (adjusted IRR 4.34, CI: 3.94-4.78) groups. Similar patterns were observed in risk of non-fatal MI or non-fatal ischaemic stroke (**Table S5**). When modelled as a continuous variable, rising BMI had a steeper relationship with cardiovascular mortality in people without DM, although with overlapping 95% confidence intervals (**Figure S1**).

**Table 4.** Unadjusted and adjusted incidence rate ratios (IRR) for all-cause and cardiovascular mortality obtained from multivariable Poisson regression analysis in participants grouped by diabetes and ethnicity adjusted BMI category. *Adjusted for age, sex, ethnicity, and smoking status. Abbreviations: body mass index (BMI); confidence interval (CI); incident rate ratio (IRR).

| Comparison | All-cause mortality |  | Cardiovascular mortality |  |
| --- | --- | --- | --- | --- |
|  | Unadjusted IRR (95% CI) | Adjusted IRR* (95% CI) | Unadjusted IRR (95% CI) | Adjusted IRR* (95% CI) |
| Normal + non-diabetes | 1.00 (reference) | 1.00 (reference) | 1.00 (reference) | 1.00 (reference) |
| Overweight + non-diabetes | 1.15 (1.11-1.18, p <0.001) | 0.97 (0.94-1.00, p = 0.063) | 1.38 (1.28-1.48, p <0.001) | 1.09 (1.02-1.18, p=0.015) |
| Obese + non-diabetes | 1.38 (1.34-1.43, p <0.001) | 1.25 (1.21-1.28, p <0.001) | 1.96 (1.82-2.12, p<0.001) | 1.70 (1.58-1.84, p<0.001) |
| Normal + diabetes | 3.16 (2.84-3.50, p <0.001) | 2.00 (1.80-2.23, p <0.001) | 4.38 (3.47-5.23, p <0.001) | 2.45 (1.94-3.10, p<0.001) |
| Overweight + diabetes | 3.01 (2.83-3.20, p <0.001) | 1.78 (1.67-1.89, p <0.001) | 5.71 (5.06-6.44, p <0.001) | 2.90 (2.56-3.28, p <0.001) |
| Obese + diabetes | 3.52 (3.36-3.69, p <0.001) | 2.40 (2.29-2.52, p <0.001) | 7.05 (6.40-7.75, p<0.001) | 4.34 (3.94-4.78, p <0.001) |

**Figure 2.**
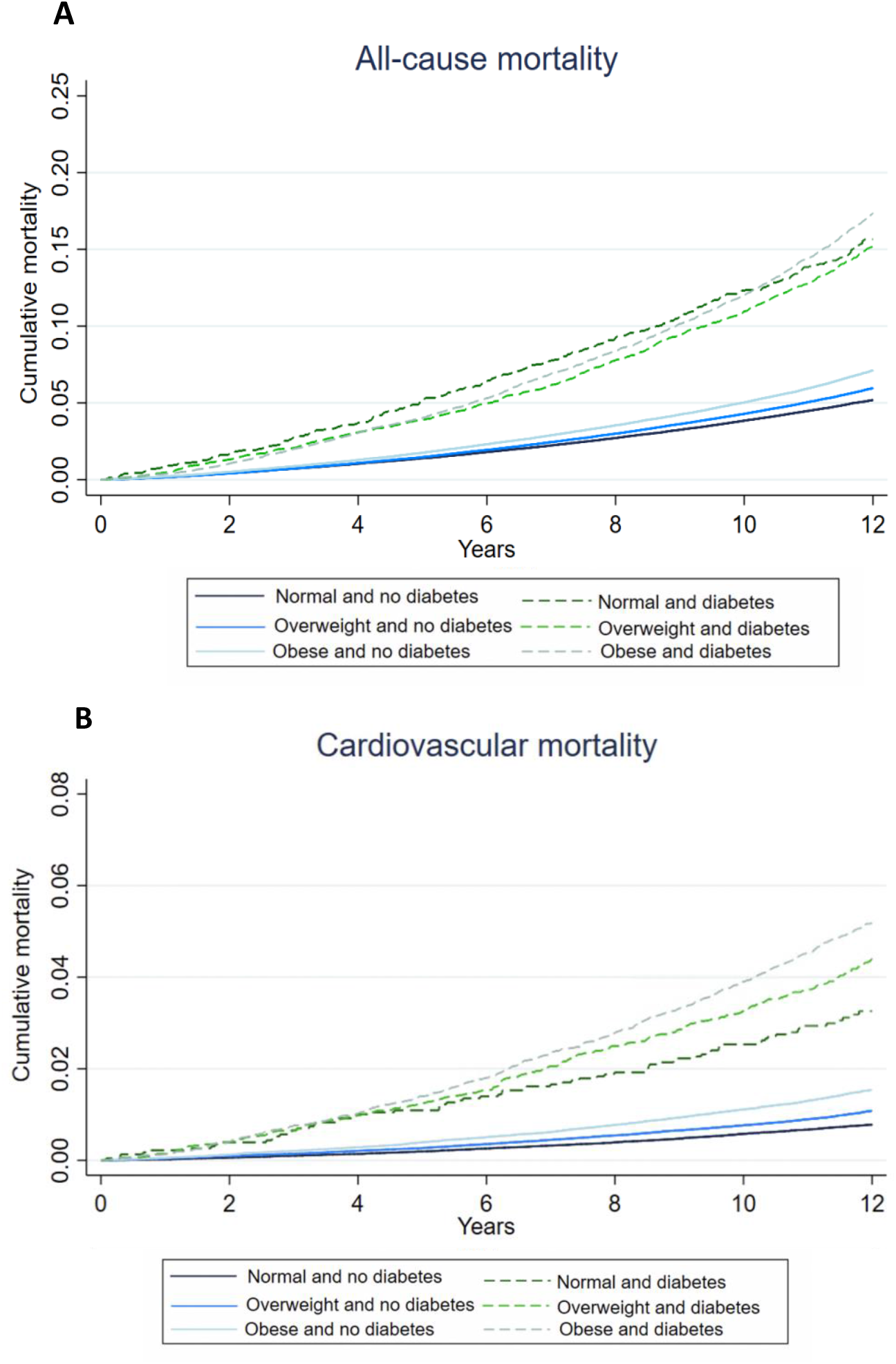
All-cause and cardiovascular mortality. Kaplan-Meier mortality curves illustrating cumulative all-cause (A) and cardiovascular (B) mortality stratified by ethnicity-adjusted BMI category and diabetes status.

## Discussion

We present a detailed analysis of cardiovascular phenotypes and outcomes in relation to metabolic parameters in a large cohort stratified according to their baseline DM status and BMI category. Cardiovascular mortality and non-fatal events were more common as BMI rose, but in the presence of DM event rates were much greater. Indeed, people with DM and normal BMI experienced substantially higher adjusted event rates than obese people without DM. Multimodality cardiovascular imaging data corroborated progressive cardiovascular abnormalities with rising BMI in people without DM. Notably, the cardiovascular imaging phenotype of DM with normal BMI was broadly comparable to obesity without DM, and there was only modest progression of these cardiovascular abnormalities with rising BMI in the DM group. Furthermore, we found that the duration of DM and HbA1c correlated poorly with cardiovascular phenotype in people with DM, with metrics of adiposity demonstrating the strongest (albeit modest) correlation. This raises the possibility that the ideal target range for metrics of adiposity may be lower than currently proposed in people with DM. However, there is also clearly scope to improve adherence to existing targets around modifiable cardiovascular risk factors in people with DM and/or elevated BMI.

### Epidemiological insights

To the best of our knowledge, our study is the largest investigation of the interaction between diabetes and obesity in terms of cardiovascular phenotype and long-term outcomes. However, it is helpful to present our findings in the context of other epidemiological studies. For example, in over 10 million participants from Asia, North America, Europe and Australasia, Di Angelantonio *et al* showed that being overweight *or* obese was associated with higher all-cause and cardiovascular mortality compared to normal weight. [13] However, they did not directly compare cardiovascular mortality in people stratified by DM status. In a prospective cohort of 10,568 people with type 2 DM of whom most did not have concomitant cardiovascular disease, Costanzo *et al* showed overweight and obese people were more likely to be hospitalized for myocardial infarction or stroke, although cardiovascular mortality was not reported. [33,34] In a recent meta-analysis of 16 cohort studies including 445 125 people with type 2 DM, Kwon *et al* report a U-shaped relationship between increasing BMI with all-cause and cardiovascular mortality with a nadir of risk at a BMI of 28–30 kg/m^2^ and 29–31 kg/m^2^ respectively. [35] Again, no comparison was made with data from people without DM.

### Cardiovascular imaging insights

We found that participants with DM had more advanced atherosclerosis (defined by CIMT) and greater arterial stiffness than participants without DM within any given BMI category. Moreover, participants with normal BMI and DM had a nominally higher CIMT than any other DM-BMI category, emphasising the substantial burden of arterial disease in this group. Participants with DM also had evidence of supraphysiological cardiac contractility without differences in cardiac output. This observation, combined with the associated increased arterial stiffness, suggests chronically elevated left ventricular afterload – which is a risk factor for incident heart failure and atrial fibrillation, amongst others. [36] Indeed, data from serial cardiac MRI studies in people with uncomplicated type 2 DM have revealed important reductions in LVEF over a 6-year period. [37] Whilst we found statistically lower LVEF in people with DM, these were smaller than the margin of error with any cardiac imaging modality. Given we observed much larger proportional differences in CCI, this may be a better biomarker of early diabetic heart disease; this warrants careful assessment in future studies.

### Clinical implications

We showed that people with DM and normal BMI had significantly elevated WHR and a numerically greater abdominal fat ratio and TAATI on abdominal MRI compared to people of normal BMI without DM. These measures are suggestive of unfavourable body composition, and greater visceral as opposed to subcutaneous adipose deposition. Chowdary *et al* recently described that people with diabetes have greater visceral adiposity (including epicardial adiposity) than those without diabetes even when of normal weight. [15] Visceral adiposity is independently associated with higher 10-year cardiovascular disease risk. [38] It is notable that indices of adiposity were the strongest (albeit modest) correlates of abnormal cardiovascular phenotypes in people with and without DM in our analysis. Hence, better assessment of visceral adiposity in routine practice may allow clinicians to define high risk groups and our data raise the question of whether lower BMI targets, or possibly alternate adiposity metrics, should guide use of existing and novel therapies in people with DM.

Increased risk of cardiovascular disease and death among people with DM and normal BMI is also likely to be contributed to by suboptimal control of ‘traditional’ cardiovascular risk factors. In our study, whilst this group had relatively good glycaemic control and ‘normal’ BMI, they had: low physical activity; higher than ideal systolic blood pressure; suboptimal LDL cholesterol; and approximately 1 in 7 currently smoked. These data emphasise the potential benefits of more effective screening and application of existing cardiovascular risk modification guidelines. Nevertheless, the other groups in our analysis also had suboptimal cardiovascular risk factor profiles, emphasising the challenges of preventative medicine. Interestingly, systemic inflammation (as measured by serum CRP) was not notably different between people with and without DM of normal BMI, which conflicts with some published data. [39] However, systemic inflammation did increase with greater obesity irrespective of DM status – a well-documented phenomenon. [40] The modest correlation of all tested cardiovascular risk factors with cardiovascular imaging phenotypes in people with diabetes, also supports the need to define better routine clinical biomarkers of cardiovascular disease phenotypes in people with diabetes.

### Strengths and limitations

Our study has several strengths including: detailed cardiometabolic phenotyping with multi-modality assessment; reported outcomes on all-cause mortality, cardiovascular mortality, incident myocardial infarction and stroke; long follow up and statistical power to study DM with normal BMI. However, we must also acknowledge limitations of our work. First, data for every phenotypic measure of cardiometabolic disease was not available for every participant, predominantly due to the design of UKB, which only perform more complex assessments in a subset of participants. Moreover, there is a survivor bias in participants who underwent detailed imaging assessment as this only commenced in 2014. Imaged participants were less obese, had lower smoking prevalence, greater educational attainment and more likely to be white ethnicity. [19] Second, we did not stratify by type of DM since only 404 participants had a self-reported diagnosis of type 1 DM. Third, our work is observational in nature so causality cannot be inferred in the associations we found. Fourth, participants were recruited before the use of agents such as sodium-glucose cotransporter 2 (SGLT2) inhibitors, which reduce incident cardiovascular disease and improve survival in patients with diabetes. [41] Therefore, our observed event rates in people with DM may be higher than contemporary rates. Equally, UKB is not representative of the whole UK population regarding socioeconomic deprivation (SED), some non-communicable diseases and ethnic minorities.[42] Whilst this means caution should be applied in extrapolating observed event rates to the UK population, UKB remains a robust resource to define exposure-disease relationships. [42]

## Conclusion

Both obesity and DM are independently associated with more advanced cardiovascular disease and more frequent major adverse cardiovascular events. Whilst adiposity metrics are more strongly correlated with cardiovascular biomarkers than diabetes-oriented metrics, both correlate weakly, suggesting other factors underpin the high cardiovascular risk of normal-weight diabetes. Whilst there is clear scope for better use of existing screening and preventative approaches in people with DM, our data suggest that more refined risk assessment aligned with targeted preventative interventions are needed to improve outcomes.

## Data Availability

The UK Biobank resource is open to all bona fide researchers.

https://www.ukbiobank.ac.uk/

## Acknowledgements

This research has been conducted using the UK Biobank resource - application number 59585. This work uses data provided by patients and collected by the NHS as part of their care and support; Copyright © (2022), NHS Digital; Re-used with the permission of UK Biobank. All rights reserved. This research used data assets made available by National Safe Haven as part of the Data and Connectivity National Core Study, led by Health Data Research UK in partnership with the Office for National Statistics and funded by UK Research and Innovation (research which commenced between 1st October 2020 – 31st March 2021 grant ref MC_PC_20029; 1st April 2021 -30th September 2022 grant ref MC_PC_20058). MD was supported by a British Heart Foundation Clinical Research Training Fellowship. SS is supported by a British Heart Foundation Clinical Research Training Fellowship. EL is funded by Wellcome Trust Clinical Career Development Fellowship (221690/Z/20/Z) and receives support from National Institute for Health and Care Research (NIHR) Leeds Biomedical Research Centre. The views expressed are those of the author(s) and not necessarily those of the NHS, the NIHR or the Department of Health and Social Care. LDR was supported by the Diabetes UK RD Lawrence Fellowship (16/0005382). KJG is a National Institute for Health and Care Research Academic Clinical Lecturer. MAB is supported by a British Heart Foundation Intermediate Clinical Research Fellowship (FS/18/12/33270). MTK is a British Heart Foundation professor. RMC was supported by a British Heart Foundation Intermediate Clinical Research Fellowship.

## Transparency Statement

RC affirms that the manuscript is an honest, accurate, and transparent account of the study being reported; no important aspects of the study have been omitted; there are no discrepancies from the study as originally planned.

## Data sharing Statement

The UK Biobank resource is open to all *bona fide* researchers.

## Authorship Statement

Guarantors – MD and RMC; Study conception – all authors; Data analysis – OB and MD; Manuscript drafting – OB, MD, HM and RC; Critical revision of manuscript: MG, MCR, JG, SS, SBW, KP, LR, EL, RA, KJG, MAB, MTK.

## References

1 GBD 2015 Risk Factors Collaborators. Global, regional, and national comparative risk assessment of 79 behavioural, environmental and occupational, and metabolic risks or clusters of risks, 1990-2015: a systematic analysis for the Global Burden of Disease Study 2015. Lancet 2016;388:1659–724. doi:10.1016/S0140-6736(16)31679-8

2 Bogers RP, Bemelmans WJE, Hoogenveen RT, et al. Association of overweight with increased risk of coronary heart disease partly independent of blood pressure and cholesterol levels: a meta-analysis of 21 cohort studies including more than 300 000 persons. Arch Intern Med 2007;167:1720–8. doi:10.1001/archinte.167.16.1720

3 Adams KF, Schatzkin A, Harris TB, et al. Overweight, obesity, and mortality in a large prospective cohort of persons 50 to 71 years old. N Engl J Med 2006;355:763–78. doi:10.1056/NEJMoa055643

4 Loos RJF, Yeo GSH. The genetics of obesity: from discovery to biology. Nat Rev Genet 2022;23:120–33. doi:10.1038/s41576-021-00414-z

5 Ference B. Integrating the Effect of BMI and Polygenic Scores to estimate Lifetime Risk and Identify Optimal Treatment Targets to Prevent or Reverse Diabetes. In: ESC Congress - The Digital Experience. 2020.

6 Hjerkind KV, Stenehjem JS, Nilsen TIL. Adiposity, physical activity and risk of diabetes mellitus: prospective data from the population-based HUNT study, Norway. BMJ Open 2017;7:e013142. doi:10.1136/bmjopen-2016-013142

7 Gatineau M, Hancock C, Holman N, et al. Adult obesity and type 2 diabetes. Oxford: Public Health England 2014.

8 Chait A, Bornfeldt KE. Diabetes and atherosclerosis: is there a role for hyperglycemia? J Lipid Res 2009;50 Suppl:S335–9. doi:10.1194/jlr.R800059-JLR200

9 Wildman RP, Mackey RH, Bostom A, et al. Measures of obesity are associated with vascular stiffness in young and older adults. Hypertension 2003;42:468–73. doi:10.1161/01.HYP.0000090360.78539.CD

10 Brown OI, Bridge KI, Kearney MT. Nicotinamide Adenine Dinucleotide Phosphate Oxidases in Glucose Homeostasis and Diabetes-Related Endothelial Cell Dysfunction. Cells 2021;10. doi:10.3390/cells10092315

11 Raghavan S, Vassy JL, Ho Y-L, et al. Diabetes Mellitus-Related All-Cause and Cardiovascular Mortality in a National Cohort of Adults. J Am Heart Assoc 2019;8:e011295. doi:10.1161/JAHA.118.011295

12 Chen Y, Copeland WK, Vedanthan R, et al. Association between body mass index and cardiovascular disease mortality in east Asians and south Asians: pooled analysis of prospective data from the Asia Cohort Consortium. BMJ 2013;347:f5446. doi:10.1136/bmj.f5446

13 Global BMI Mortality Collaboration, di Angelantonio E, Bhupathiraju S, et al. Body-mass index and all-cause mortality: individual-participant-data meta-analysis of 239 prospective studies in four continents. Lancet 2016;388:776–86. doi:10.1016/S0140-6736(16)30175-1

14 Levelt E, Pavlides M, Banerjee R, et al. Ectopic and Visceral Fat Deposition in Lean and Obese Patients With Type 2 Diabetes. J Am Coll Cardiol 2016;68:53–63. doi:10.1016/j.jacc.2016.03.597

15 Chowdhary A, Thirunavukarasu S, Jex N, et al. Coronary microvascular function and visceral adiposity in patients with normal body weight and type 2 diabetes. Obesity (Silver Spring) 2022;30:1079–90. doi:10.1002/oby.23413

16 Sudlow C, Gallacher J, Allen N, et al. UK biobank: an open access resource for identifying the causes of a wide range of complex diseases of middle and old age. PLoS Med 2015;12:e1001779. doi:10.1371/journal.pmed.1001779

17 WHO Expert Consultation. Appropriate body-mass index for Asian populations and its implications for policy and intervention strategies. Lancet 2004;363:157–63. doi:10.1016/S0140-6736(03)15268-3

18 Drozd M, Pujades-Rodriguez M, Lillie PJ, et al. Non-communicable disease, sociodemographic factors, and risk of death from infection: a UK Biobank observational cohort study. Lancet Infect Dis 2021;21:1184–91. doi:10.1016/S1473-3099(20)30978-6

19 Littlejohns TJ, Holliday J, Gibson LM, et al. The UK Biobank imaging enhancement of 100,000 participants: rationale, data collection, management and future directions. Nat Commun. 2020;11. doi:10.1038/s41467-020-15948-9

20 Raisi-Estabragh Z, Harvey NC, Neubauer S, et al. Cardiovascular magnetic resonance imaging in the UK Biobank: a major international health research resource. Eur Heart J Cardiovasc Imaging 2021;22:251–8. doi:10.1093/ehjci/jeaa297

21 Coffey S, Lewandowski AJ, Garratt S, et al. Protocol and quality assurance for carotid imaging in 100,000 participants of UK Biobank: development and assessment. Eur J Prev Cardiol 2017;24:1799–806. doi:10.1177/2047487317732273

22 Zekavat SM, Aragam K, Emdin C, et al. Genetic Association of Finger Photoplethysmography-Derived Arterial Stiffness Index With Blood Pressure and Coronary Artery Disease. Arterioscler Thromb Vasc Biol 2019;39:1253–61. doi:10.1161/ATVBAHA.119.312626

23 Liu Y, Basty N, Whitcher B, et al. Genetic architecture of 11 organ traits derived from abdominal MRI using deep learning. Elife 2021;10. doi:10.7554/eLife.65554

24 Tong TY, Key TJ, Sobiecki JG, et al. Anthropometric and physiologic characteristics in white and British Indian vegetarians and nonvegetarians in the UK Biobank. Am J Clin Nutr 2018;107:909–20. doi:10.1093/ajcn/nqy042

25 Elliott P, Peakman TC, UK Biobank. The UK Biobank sample handling and storage protocol for the collection, processing and archiving of human blood and urine. Int J Epidemiol 2008;37:234–44. doi:10.1093/ije/dym276

26 Gierula J, Paton MF, Lowry JE, et al. Rate-Response Programming Tailored to the Force-Frequency Relationship Improves Exercise Tolerance in Chronic Heart Failure. JACC Heart Fail 2018;6:105–13. doi:10.1016/j.jchf.2017.09.018

27 Bombardini T, Correia MJ, Cicerone C, et al. Force-frequency relationship in the echocardiography laboratory: a noninvasive assessment of Bowditch treppe? J Am Soc Echocardiogr 2003;16:646–55. doi:10.1016/s0894-7317(03)00221-9

28 Straw S, Cole C, Brown OI, et al. Cardiac contractility index identifies systolic dysfunction in preserved ejection fraction heart failure. medRxiv Published Online First: 2022. doi:doi: 10.1101/2022.11.22.22282605

29 Linge J, Borga M, West J, et al. Body Composition Profiling in the UK Biobank Imaging Study. Obesity (Silver Spring) 2018;26:1785–95. doi:10.1002/oby.22210

30 Drozd M, Pujades-Rodriguez M, Sun F, et al. Causes of Death in People With Cardiovascular Disease: A UK Biobank Cohort Study. J Am Heart Assoc 2021;10:e023188. doi:10.1161/JAHA.121.023188

31 Schnier C BKNJSC. Definitions of Acute Myocardial Infarction and Main Myocardial Infarction Pathological Types UK Biobank Phase 1 Outcomes Adjudication.

32 Millett ERC, Peters SAE, Woodward M. Sex differences in risk factors for myocardial infarction: cohort study of UK Biobank participants. BMJ 2018;363:k4247. doi:10.1136/bmj.k4247

33 Brown O, Costanzo P, Clark AL, et al. Relationship between a single measurement at baseline of body mass index, glycated hemoglobin, and the risk of mortality and cardiovascular morbidity in type 2 diabetes mellitus. Cardiovasc Endocrinol Metab 2020;9:177–82. doi:10.1097/XCE.0000000000000202

34 Costanzo P, Cleland JGF, Pellicori P, et al. The obesity paradox in type 2 diabetes mellitus: Relationship of body mass index to prognosis a cohort study. Ann Intern Med 2015;162:610–8. doi:10.7326/M14-1551

35 Kwon Y, Kim HJ, Park S, et al. Body Mass Index-Related Mortality in Patients with Type 2 Diabetes and Heterogeneity in Obesity Paradox Studies: A Dose-Response Meta-Analysis. PLoS One 2017;12:e0168247. doi:10.1371/journal.pone.0168247

36 Fuchs FD, Whelton PK. High Blood Pressure and Cardiovascular Disease. Hypertension 2020;75:285–92. doi:10.1161/HYPERTENSIONAHA.119.14240

37 Chowdhary A, Jex N, Thirunavukarasu S, et al. Prospective Longitudinal Characterization of the Relationship between Diabetes and Cardiac Structural and Functional Changes. Cardiol Res Pract 2022;2022:6401180. doi:10.1155/2022/6401180

38 Kouli G-M, Panagiotakos DB, Kyrou I, et al. Visceral adiposity index and 10-year cardiovascular disease incidence: The ATTICA study. Nutr Metab Cardiovasc Dis 2017;27:881–9. doi:10.1016/j.numecd.2017.06.015

39 Donath MY, Shoelson SE. Type 2 diabetes as an inflammatory disease. Nat Rev Immunol 2011;11:98–107. doi:10.1038/nri2925

40 Ellulu MS, Patimah I, Khaza’ai H, et al. Obesity and inflammation: the linking mechanism and the complications. Arch Med Sci 2017;13:851–63. doi:10.5114/aoms.2016.58928

41 McGuire DK, Shih WJ, Cosentino F, et al. Association of SGLT2 Inhibitors With Cardiovascular and Kidney Outcomes in Patients With Type 2 Diabetes: A Meta-analysis. JAMA Cardiol 2021;6:148–58. doi:10.1001/jamacardio.2020.4511

42 Fry A, Littlejohns TJ, Sudlow C, et al. Comparison of Sociodemographic and Health-Related Characteristics of UK Biobank Participants With Those of the General Population. Am J Epidemiol 2017;186:1026–34. doi:10.1093/aje/kwx246

