## Supplementary material for "The relationship between diabetes, obesity and cardiovascular disease phenotypes: a UK Biobank cohort study"

| | | No Diabetes | | | Diabetes | | |
|---|---|---|---|---|---|---|---|
| | Missing | Normal (N=143 847) | Overweight (N=185 758) | Obese (N=99 589) | Normal (N=2290) | Overweight (N=7732) | Obese (N=12 429) |
| | | | | *Cardiometabolic medication* | | | |
| **Angiotensin converting enzyme inhibitor, *n (%)*** | 0 | 5482 (3.8)*† | 13 125 (7.1)*† | 11 430 (11.5)*† | 621 (27.1)*† | 2564 (33.2)*† | 4701 (37.8)*† |
| **Angiotensin receptor blocker, *n (%)*** | 0 | 2294 (1.6)*† | 6451 (3.5)*† | 6628 (6.7)*† | 213 (9.3)*† | 1093 (14.1)*† | 2418 (19.5)*† |
| **Beta-blocker, *n (%)*** | 0 | 4914 (3.4)*† | 11 596 (6.2)*† | 9651 (9.7)*† | 251 (11.0)*† | 1357 (17.6)*† | 2873 (23.1)*† |
| **Calcium channel blocker, *n (%)*** | 0 | 4864 (3.4)*† | 11 868 (6.4)*† | 10 415 (10.5)*† | 346 (15.1)*† | 1715 (22.2)*† | 3441 (27.7)*† |
| **Statin, *n (%)*** | 0 | 10 156 (7.1)*† | 25 703 (13.8)*† | 18 269 (18.3)*† | 1425 (62.2)*† | 5393 (69.8)*† | 8927 (71.8)*† |
| **Aspirin, *n (%)*** | 0 | 11 449 (8.0)*† | 22 765 (12.3)*† | 15 577 (15.6)*† | 995 (43.5)*† | 3974 (51.4)*† | 6642 (53.4)*† |
| **Loop diuretic, *n (%)*** | 0 | 346 (0.2)*† | 888 (0.5)*† | 1713 (1.7)*† | 65 (2.8)*† | 241 (3.1)*† | 957 (7.7)*† |
| **Thiazide diuretic, *n (%)*** | 0 | 4641 (3.2)*† | 11 890 (6.4)*† | 11 337 (11.4)*† | 201 (8.8)*† | 1146 (14.8)*† | 2591 (20.9)*† |
| | | | | *Diabetes medication* | | | |
| **Insulin, *n (%)*** | 0 | <50 (0.0)*† | <50 (0.0)*† | <50 (0.0)*† | 811 (35.4)*† | 1501 (19.4)*† | 2227 (17.9)*† |
| **Metformin, *n (%)*** | 0 | <50 (0.0)*† | 54 (0.0)*† | 86 (0.1)*† | 861 (37.6)*† | 4143 (53.6)*† | 7968 (64.1)*† |
| **Sulphonylurea , *n (%)*** | 0 | <50 (0.0)† | <50 (0.0)† | (0.0)† | 396 (17.3)*† | 1682 (21.8)*† | 2824 (22.7)*† |
| **Thiazolidinediones, *n (%)*** | 0 | <50 (0.0)*† | <50 (0.0)*† | <50 (0.0)*† | 80 (3.5)*† | 500 (6.5)*† | 1525 (12.3)*† |
| **Meglitinides, *n (%)*** | 0 | <50 (0.0)† | <50 (0.0)† | <50 (0.0)† | <50 (0.5)† | <50 (0.5)† | 71 (0.6)† |

**Table S1** - **Participant medications at study recruitment**. Participants stratified by diabetes status and then by ethnicity adjusted BMI category. Normal: BMI ≥18.5kg/m$^2$ to <25 kg/m$^2$ *or* ≥18.5kg/m$^2$ to <23 kg/m$^2$ if south Asian ethnicity; Overweight: ≥25kg/m$^2$ to <30 kg/m$^2$ *or* ≥23kg/m$^2$ to <27.5 kg/m$^2$ if south Asian ethnicity; Obese: ≥30kg/m$^2$ *or* ≥27.5kg/m$^2$ if south Asian ethnicity. Categorical data presented as n (%). * represents chi$^2$ test ≤ 0.05 between BMI categories for categorical variables respectively within diabetes or non-diabetes groups. † represents p value <0.05 between each BMI category in diabetes participants and their respective BMI category in non-diabetes participants from chi$^2$ test for categorical variables. Where fewer than 50 participants are within any group, UK Biobank requires that the specific number of participants is not listed to reduce the risk of de-anonymisation.

| | No Diabetes | | | Diabetes | | |
|---|---|---|---|---|---|---|
| | Normal | Overweight | Obese | Normal | Overweight | Obese |
| ***Male*** | | | | | | |
| *Diabetes metrics* | | | | | | |
| **Diabetes duration, *years*** | - | - | - | 8 (3-18)* | 6 (2-11)* | 6 (2-10)* |
| **Age of diabetes diagnosis, *years*** | - | - | - | 52 (39-59)* | 55 (46-60)* | 53 (46-59)* |
| *Body composition* | | | | | | |
| **BMI** | 23.5 (22.3-24.3)*† | 27.2 (26.1-28.5)*† | 31.1 (30.9-34.3)*† | 23.7 (22.5)*† | 27.7 (26.4-28.9)*† | 33.5 (31.5-36.5)*† |
| **Waist circumference, cm** | 86 (82-90)*† | 96 (92-100)*† | 108 (103-114)*† | 88 (83-91)*† | 98 (93-102)*† | 113 (106-120)*† |
| **Hip circumference, cm** | 97 (94-100)*† | 103 (100-106)*† | 110 (106-114)*† | 96 (93-99)*† | 102 (99-105)*† | 111 (107-117)*† |
| **Waist to hip ratio** | 0.88 (0.85-0.92)*† | 0.93 (0.90-0.96)*† | 0.98 (0.95-1.02)*† | 0.91 (0.87-0.95)*† | 0.96 (0.92-0.99)*† | 1.01 (0.97-1.05)*† |
| **Body fat percentage** | 19.7 (16.6-22.5)*† | 25.1 (22.6-27.6)*† | 30.8 (28.3-33.4)*† | 20.6 (17.2-23.5)*† | 26.3 (23.7-28.7)*† | 32.4 (29.6-35.5)*† |
| **Whole body fat mass, *kg*** | 14.3 (11.7-16.7)*† | 21.1 (18.4-23.8)*† | 30.4 (27.0-34.9)*† | 14.7 (12.0-17.0)*† | 21.9 (19.1-24.7)*† | 32.9 (28.5-39.1)*† |
| **Whole body impedance, *ohms*** | 580 (547-615)* | 536 (507-566)* | 489 (460-519)*† | 578 (542-617)* | 535 (503-569)* | 478 (446-513)*† |
| *Lipids* | | | | | | |
| **Serum Apolipoprotein A, *g/L*** | 1.5 (1.3-1.6)*† | 1.4 (1.3-1.6)*† | 1.3 (1.2-1.5)*† | 1.4 (1.3-1.6)*† | 1.3 (1.2-1.5)*† | 1.3 (1.2-1.4)*† |
| **Serum Apolipoprotein B, *g/L*** | 1.0 (0.8-1.1)*† | 1.0 (0.9-1.2)*† | 1.0 (0.9-1.2)*† | 0.8 (0.7-0.9)*† | 0.8 (0.7-0.9)*† | 0.8 (0.7-1.0)*† |
| **Serum total cholesterol, *mmol/L*** | 5.5 (4.8-6.2)*† | 5.6 (4.9-6.3)*† | 5.5 (4.7-6.3)*† | 4.2 (3.7-4.9)*† | 4.2 (3.7-4.9)*† | 4.2 (3.6-4.8)*† |
| **Serum HDL cholesterol, *mmol/L*** | 1.4 (1.2-1.6)*† | 1.2 (1.1-1.4)*† | 1.1 (1.0-1.3)*† | 1.3 (1.1-1.5)*† | 1.1 (1.0-1.3)*† | 1.0 (0.9-1.2)*† |
| **Serum LDL cholesterol, *mmol/L*** | 3.4 (2.9-4.0)*† | 3.6 (3.0-4.1)*† | 3.5 (2.9-4.1)*† | 2.4 (2.1-2.9)*† | 2.5 (2.1-3.0)*† | 2.5 (2.1-3.0)*† |
| **Serum lipoprotein A, *nmol/L*** | 20.2 (9.4-61.6)*† | 20.2 (9.3-61.0)* | 18.7 (8.7-63.1)*† | 18.7 (8.4-58.6)*† | 20.2 (8.8-67.3)* | 16.7 (7.8-62.0)*† |
| **Serum triglycerides, *mmol/L*** | 1.3 (0.9-1.8)*† | 1.7 (1.2-2.5)* | 2.1 (1.5-2.9)* | 1.3 (0.9-1.8)*† | 1.7 (1.2-2.5)* | 2.1 (1.5-2.9)* |
| *Biochemistry* | | | | | | |
| **Creatinine, *μmol/L*** | 78 (71-86)*† | 81 (74-89)*† | 81 (73-90)*† | 76 (68-86)*† | 78 (70-88)*† | 79 (69-90)*† |
| **Serum cystatin C, *mg/L*** | 0.88 (0.81-0.96)*† | 0.91 (0.84-1.00)*† | 0.96 (0.88-1.06)*† | 0.90 (0.81-1.02)*† | 0.93 (0.84-1.04)*† | 0.98 (0.88-1.11)*† |
| **Urinary microalbumin, *mg/L*** | 10.6 (8.1-17.2)*† | 11.1 (8.3-18.6)*† | 12.7 (8.8-23.3)* | 15.1 (9.4-33.4)*† | 15.4(9.9-34.7)*† | 19.8 (10.9-51.6)* |
| **Alanine aminotransferase, *U/L*** | 20 (16-25)*† | 24 (19-31)*† | 29 (22-39)* | 22 (17-28)*† | 25 (19-33)*† | 29 (21-40)* |
| **C-reactive protein, *mg/L*** | 0.8 (0.4-1.6)*† | 1.2 (0.7-2.3)*† | 2.0 (1.1-3.6)*† | 0.8 (0.4-1.8)*† | 1.2 (0.6-2.3)*† | 2.0 (1.1-3.8)*† |
| *Diabetes related biomarkers* | | | | | | |
| **Glucose, *mmol/L*** | 4.9 (4.5-5.2)*† | 4.9 (4.6-5.3)*† | 5.0 (4.6-5.4)*† | 6.5 (5.2-9.5)*† | 6.5 (5.3-8.7)*† | 6.7 (5.4-9.2)*† |
| **HbA1c, *mmol/mol*** | 34 (32-37)*† | 35 (33-37)*† | 36 (34-39)*† | 49 (42-58)*† | 50 (43-58)*† | 51 (44-61)*† |
| **IGF-1, *nmol/L*** | 22.0 (18.6-25.3)*† | 22.2 (18.9-25.6)*† | 21.0 (17.3-24.5)*† | 21.3 (17.4-25.5)*† | 21.2(17.1-25.0)*† | 19.1(14.9-23.4)*† |
| *Abdominal MRI* | | | | | | |
| **Abdominal fat ratio, *fraction*** | 0.37 (0.31-0.43)* | 0.46 (0.41-0.51)* | 0.54 (0.48-0.58)*† | 0.42 (0.33-0.48)* | 0.47 (0.44-0.54)* | 0.57 (0.54-0.61)*† |
| **Total abdominal adipose tissue index, *$L/m^2$*** | 2.2 (1.7-2.8)* | 3.5 (2.9-4.1)*† | 5.1 (4.3-5.9)*† | 2.5 (1.8-3.2)* | 3.7 (3.0-4.4)*† | 5.6 (5.1-6.4)*† |
| ***Female*** | | | | | | |
| *Diabetes metrics* | | | | | | |
| **Diabetes duration, *years*** | - | - | - | 7 (3-21)* | 5 (2-12)* | 5 (2-10)* |
| **Age of diabetes diagnosis, *years*** | - | - | - | 52 (34-60)* | 55 (45-60)* | 54 (45-59)* |

| | | | | | | |
|---|---|---|---|---|---|---|
| **BMI** | 22.9 (21.6-24.0)*† | 27.0 (25.9-28.3)*† | 33.0 (31.2-35.9)*† | 23.1 (21.9-24.1)*† | 27.6 (26.3-28.8)*† | 34.9 (32.2-38.9)*† |
| **Waist circumference, cm** | 74 (70-79)*† | 85 (80-90)*† | 99 (93-105)*† | 77 (72-82)*† | 90 (85-94)*† | 106 (99-114)*† |
| **Hip circumference, cm** | 96 (92-99)*† | 103 (100-107)*† | 115 (110-121)*† | 95 (91-98)*† | 102 (98-106)*† | 117 (110-125)*† |
| **Waist to hip ratio** | 0.78 (0.74-0.82)*† | 0.82 (0.78-0.87)*† | 0.86 (0.82-0.90)*† | 0.81 (0.76-0.87)*† | 0.88 (0.83-0.93)*† | 0.90 (0.86-0.95)*† |
| **Body fat percentage** | 31.1 (27.8-34.1)* | 38.0 (35.6-40.3)* | 44.3 (41.8-46.8)*† | 31.2 (27.6-34.3)* | 38.1 (35.4-40.4)* | 45.1 (42.3-48.0)*† |
| **Whole body fat mass, *kg*** | 18.9 (16.0-21.6)* | 27.0 (24.2-30.1)* | 38.3 (34.1-43.9)*† | 19.0 (15.6-21.7)* | 27.1 (23.9-30.2)* | 41.0 (35.1-48.2)*† |
| **Whole body impedance, *ohms*** | 690 (650-732)*† | 649 (612-688)*† | 590 (630-667)*† | 679 (636-721)*† | 635 (595-677)*† | 564 (520-608)*† |
| *Lipids* | | | | | | |
| **Serum Apolipoprotein A, *g/L*** | 1.7 (1.5-1.9)*† | 1.6 (1.4-1.8)*† | 1.5 (1.4-1.7)*† | 1.6 (1.4-1.8)*† | 1.5 (1.4-1.7)*† | 1.4 (1.3-1.6)*† |
| **Serum Apolipoprotein B, *g/L*** | 1.0 (0.8-1.1)*† | 1.1 (0.9-1.2)*† | 1.1 (0.9-1.2)*† | 0.8 (0.7-0.9)*† | 0.8 (0.7-1.0)*† | 0.9 (0.7-1.0)*† |
| **Serum total cholesterol, *mmol/L*** | 5.8 (5.1-6.5)*† | 6.0 (5.2-6.7)*† | 5.9 (5.1-6.6)*† | 4.7 (4.1-5.4)*† | 4.7 (4.0-5.3)*† | 4.6 (4.0-5.2)*† |
| **Serum HDL cholesterol, *mmol/L*** | 1.7 (1.5-2.0)*† | 1.5 (1.3-1.8)*† | 1.4 (1.2-1.6)*† | 1.6 (1.3-1.9)*† | 1.3 (1.1-1.6)*† | 1.2 (1.1-1.4)*† |
| **Serum LDL cholesterol, *mmol/L*** | 3.5 (2.9-4.0)*† | 3.7 (3.1-4.3)*† | 3.7 (3.1-4.3)*† | 2.6 (2.2-3.1)*† | 2.7 (2.2-3.2)*† | 2.7 (2.3-3.2)*† |
| **Serum lipoprotein A, *nmol/L*** | 20.8 (9.5-59.7)* | 22.9 (10.2-62.1)* | 24.0 (10.4-65.4)*† | 21.4 (9.7-63.8) | 23.7 (9.6-67.2) | 20.9 (9.2-62.5)† |
| **Serum triglycerides, *mmol/L*** | 1.1 (0.8-1.5)* | 1.4 (1.0-2.0)*† | 1.7 (1.2-2.3)*† | 1.1 (0.8-1.7)* | 1.7 (1.1-2.4)*† | 1.9 (1.4-2.6)*† |
| *Biochemistry* | | | | | | |
| **Creatinine, *µmol/L*** | 62 (56-69)*† | 64 (58-71)*† | 64 (58-72)*† | 60 (54-68)*† | 62 (55-70)*† | 62 (55-72)*† |
| **Serum cystatin C, *mg/L*** | 0.82 (0.75-0.89)*† | 0.86 (0.79-0.95)*† | 0.92 (0.84-1.03)*† | 0.84 (0.76-0.94)*† | 0.90 (0.80-1.02)*† | 0.96 (0.85-1.09)*† |
| **Urinary microalbumin, *mg/L*** | 10.7 (8.2-16.5)*† | 10.7 (8.2-16.5)*† | 11.7 (8.6-19.2)*† | 13.5 (9.1-24.4)† | 13.7 (9.6-26.3)† | 14.6 (9.5-29.2)† |
| **Alanine aminotransferase, *U/L*** | 16 (13-20)*† | 18 (14-23)*† | 21 (16-27)*† | 19 (15-25)*† | 21 (16-29)*† | 23 (17-32)*† |
| **C-reactive protein, *mg/L*** | 0.8 (0.4-1.5)*† | 1.5 (0.8-2.8)*† | 3.1 (1.7-5.7)*† | 0.9 (0.5-2.0)*† | 1.6 (0.8-3.3)*† | 3.3 (1.7-6.5)*† |
| *Diabetes related biomarkers* | | | | | | |
| **Glucose, *mmol/L*** | 4.8 (4.5-5.2)*† | 4.9 (4.6-5.3)*† | 5.0 (4.7-5.4)*† | 6.3 (5.1-9.4)*† | 6.3 (5.2-8.4)*† | 6.4 (5.2-8.7)*† |
| **HbA1c, *mmol/mol*** | 34 (32-37)*† | 35 (33-37)*† | 36 (34-39)*† | 51 (44-60)*† | 50 (43-59)*† | 51 (45-60)*† |
| **IGF-1, *nmol/L*** | 21.5 (17.8-25.1)*† | 21.0 (17.3-24.7)*† | 19.2 (15.5-23.2)*† | 20.4 (16.4-24.7)† | 20.1 (16.0-24.2)† | 17.6 (13.7-21.8)† |
| *Abdominal MRI* | | | | | | |
| **Abdominal fat ratio, *fraction*** | *0.48 (0.41-0.54)** | 0.58 (0.53-0.62)* | 0.65 (0.62-0.68)*† | 0.49 (0.39-0.54)* | 0.60 (0.55-0.62)* | 0.68 (0.65-0.71)*† |
| **Total abdominal adipose tissue index, $L/m^2$** | 2.7 (2.1-3.4)* | 4.4 (3.7-5.1)* | 6.3 (5.5-7.4)*† | 3.1 (1.9-3.8)* | 4.5 (3.8-5.2)* | 7.0 (6.0-8.3)*† |

**Table S2** – **Metabolic phenotypes of study participants stratified by sex**. Participants are stratified by diabetes status, sex and then by ethnicity adjusted BMI category. Normal: BMI ≥18.5$kg/m^2$ to <25 $kg/m^2$ *or* ≥18.5$kg/m^2$ to <23 $kg/m^2$ if south Asian ethnicity; Overweight: ≥25$kg/m^2$ to <30 $kg/m^2$ *or* ≥23$kg/m^2$ to <27.5 $kg/m^2$ if south Asian ethnicity; Obese: ≥30$kg/m^2$ *or* ≥27.5$kg/m^2$ if south Asian ethnicity. Continuous data presented as median with 25$^{th}$ and 75$^{th}$ centile. Categorical data presented as n (%). * represents independent one-way ANOVA p value or chi$^2$ test <0.05 between BMI categories for continuous variables and categorical variables respectively within diabetes or non-diabetes groups. † represents p value <0.05 between each BMI category in diabetes participants and their respective BMI category in non-diabetes participants from independent t-tests for continuous variables and chi$^2$ test for categorical variables. Total abdominal adipose tissue index is defined as VAT volume + abdominal SAT volume / body surface area. Abdominal fat ratio is defined as VAT volume + abdominal SAT volume / VAT volume + abdominal SAT volume + total thigh fat-free muscle volume. Abbreviations: body mass index (BMI); subcutaneous adipose tissue (SAT); visceral adipose tissue (VAT).

| | No Diabetes | | | Diabetes | | |
|---|---|---|---|---|---|---|
| | Normal | Overweight | Obese | Normal | Overweight | Obese |
| ***Male*** | | | | | | |
| *Vital signs* | | | | | | |
| **Systolic blood pressure, *mmHg*** | 136 (125-149)*† | 142 (130-154)*† | 145 (133-157)*† | 139 (127-154)*† | 143 (132-156)*† | 144 (133-156)*† |
| **Diastolic blood pressure, *mmHg*** | 80 (73-87)*† | 84 (77-91)*† | 87 (81-94)*† | 77 (71-84)*† | 81 (74-88)*† | 83 (76-90)*† |
| **Resting heart rate, *bpm*** | 65 (58-73)*† | 66 (60-74)*† | 70 (62-79)*† | 70 (62-80)*† | 71 (63-80)*† | 74 (65-83)*† |
| *Carotid intima-media thickness* | | | | | | |
| **Mean carotid IMT, *μm*** | 672 (597-771)*† | 698 (613-796)*† | 710 (626-814)* | 728 (645-834)† | 729 (642-834)† | 713 (646-811) |
| *Cardiac MRI* | | | | | | |
| **LVEF, *%*** | 55 (51-58)* | 55 (51-59)*† | 55 (50-58)*† | 55 (53-59)* | 54 (49-58)*† | 53 (48-57)*† |
| **LVEDV, *ml*** | 151 (131-172)*† | 154 (135-175)*† | 159 (138-183)*† | 131 (114-145)*† | 139 (116-165)*† | 144 (124-172)*† |
| **LVESV, *ml*** | 67 (57-80)*† | 69 (58-81)*† | 72 (61-86)*† | 55 (49-67)*† | 64 (52-77)*† | 68 (56-84)*† |
| **LVSV, *ml*** | 82 (71-94)*† | 84 (72-96)*† | 86 (73-100)*† | 72 (61-83)† | 73 (61-88)† | 77 (62-88)† |
| **LVEDV / BSA, $ml/m^2$** | 80 (70-90)*† | 77 (68-87)*† | 74 (65-84)*† | 71 (60-77)† | 71 (62-81)† | 68 (59-79)† |
| **LVESV / BSA, $ml/m^2$** | 36 (31-42)*† | 34 (29-40)*† | 33 (28-40)*† | 28 (26-36)† | 32 (27-38)† | 32 (26-38)† |
| **LVSV / BSA, $ml/m^2$** | 43 (38-49)*† | 42 (36-48)*† | 40 (35-47)*† | 39 (33-45)*† | 37 (31-43)*† | 35 (30-41)*† |
| **Cardiac output, $L/min^{-1}$** | 4.8 (4.2-5.5)* | 5.0 (4.4-5.7)*† | 5.3 (4.5-6.1)* | 4.6 (4.1-5.4)* | 4.8 (4.0-5.5)*† | 5.1 (4.4-5.9)* |
| **Cardiac index, $L/min^{-1}/m^2$** | 2.5 (2.2-2.9)* | 2.5 (2.2-2.8)*† | 2.5 (2.1-2.8)* | 2.5 (2.2-3.0) | 2.4 (2.1-2.8)† | 2.4 (2.1-2.8) |
| **Cardiac contractility index (SBP/LVESVi)** | 3.8 (3.1-4.6)*† | 4.1 (3.4-4.9)*† | 4.3 (3.6-5.2)*† | 4.7 (4.1-5.1)† | 4.3 (3.6-5.3)† | 4.5 (3.7-5.6)† |
| *Photoplethysmography derived arterial stiffness* | | | | | | |
| **Pulse wave arterial stiffness index** | 9.0 (7.1-11.4)*† | 9.8 (7.7-12.0)*† | 10.2 (8.4-12.0)* | 9.5 (7.5-11.7)*† | 10.3 (8.3-12.2)*† | 10.0 (8.5-11.7)* |
| ***Female*** | | | | | | |
| *Vital signs* | | | | | | |
| **Systolic blood pressure, *mmHg*** | 130 (118-145)*† | 137 (124-151)*† | 140 (128-154)*† | 138 (125-152)*† | 141 (129-154)*† | 142 (131-154)*† |
| **Diastolic blood pressure, *mmHg*** | 77 (70-84)*† | 81 (74-88)*† | 85 (78-92)*† | 75 (69-82)*† | 79 (72-85)*† | 82 (76-89)*† |
| **Resting heart rate, *bpm*** | 68 (62-75)*† | 69 (63-76)*† | 72 (65-80)*† | 73 (65-80)*† | 74 (67-83)*† | 77 (68-86)*† |
| *Carotid intima-media thickness* | | | | | | |
| **Mean carotid IMT, *μm*** | 639 (576-715)*† | 654 (590-735)* | 663 (600-744)* | 715 (631-767)† | 680 (611-742) | 684 (624-762) |
| *Cardiac MRI* | | | | | | |
| **LVEF, *%*** | 57 (54-61)* | 57 (54-61)*† | 57 (54-61)*† | 58 (56-61)* | 56 (51-58)*† | 56 (52-60)*† |
| **LVEDV, *ml*** | 117 (103-132)* | 119 (105-135)*† | 128 (112-145)*† | 108 (93-129) | 109 (97-127)† | 120 (98-136)† |
| **LVESV, *ml*** | 50 (43-58)*† | 51 (43-59)* | 54 (46-63)*† | 45 (36-55)† | 48 (41-58) | 51 (42-60)† |
| **LVSV, *ml*** | 67 (58-75)* | 68 (59-77)*† | 73 (63-83)*† | 63 (53-73)* | 60 (52-68)*† | 66 (55-75)*† |
| **LVEDV / BSA, $ml/m^2$** | 71 (63-79)*† | 68 (61-76)*† | 68 (60-75)*† | 65 (57-78)† | 64 (55-72)† | 61 (54-69)† |
| **LVESV / BSA, $ml/m^2$** | 30 (26-35)*† | 29 (25-33)*† | 29 (25-33)* | 27 (23-34)† | 28 (23-33) | 26 (22-31)† |
| **LVSV / BSA, $ml/m^2$** | 40 (36-45)* | 39 (34-44)*† | 39 (33-43)*† | 39 (33-44)* | 34 (29-39)*† | 35 (29-39)*† |
| **Cardiac output, $L/min^{-1}$** | 4.1 (3.6-4.7)* | 4.3 (3.7-4.9)* | 4.6 (4.0-5.3)* | 4.2 (3.5-4.7)* | 4.2 (3.5-4.6)* | 4.4 (3.9-5.2)* |
| **Cardiac index, $L/min^{-1}/m^2$** | 2.5 (2.2-2.8)* | 2.4 (2.1-2.8)* | 2.4 (2.1-2.7)*† | 2.6 (2.1-2.7) | 2.3 (2.1-2.6) | 2.3 (2.1-2.6)† |
| **Cardiac contractility index (SBP/LVESVi)** | 4.2 (3.5-5.1)*† | 4.6 (3.8-5.6)*† | 4.8 (4.0-5.8)*† | 5.0 (4.0-5.8)† | 5.0 (4.1-6.0)† | 5.3 (4.3-6.2)† |
| *Photoplethysmography derived arterial stiffness* | | | | | | |

| | | | | | | |
|---|---|---|---|---|---|---|
| **Pulse wave arterial stiffness index** | 7.7 (6.1-9.9)*† | 8.4 (6.3-10.6)*† | 8.9 (6.7-10.9)*† | 8.4 (6.4-10.8)*† | 9.0 (6.6-11.2)*† | 9.1 (7.0-11.1)*† |

**Table S3** – **Phenotypic measurements of cardiovascular disease stratified by sex**. Participants are stratified by diabetes status, sex and then by ethnicity adjusted BMI category. Normal: BMI ≥18.5kg/m$^2$ to <25 kg/m$^2$ *or* ≥18.5kg/m$^2$ to <23 kg/m$^2$ if south Asian ethnicity; Overweight: ≥25kg/m$^2$ to <30 kg/m$^2$ *or* ≥23kg/m$^2$ to <27.5 kg/m$^2$ if south Asian ethnicity; Obese: ≥30kg/m$^2$ *or* ≥27.5kg/m$^2$ if south Asian ethnicity. Continuous data presented as median with 25$^{th}$ and 75$^{th}$ centile. Categorical data presented as n (%). * represents independent one-way ANOVA p value or chi$^2$ test <0.05 between BMI categories for continuous variables and categorical variables respectively within diabetes or non-diabetes groups. † represents p value <0.05 between each BMI category in diabetes participants and their respective BMI category in non-diabetes participants from independent t-tests for continuous variables and chi$^2$ test for categorical variables. Total abdominal adipose tissue index is defined as VAT volume + abdominal SAT volume / body surface area. Abdominal fat ratio is defined as VAT volume + abdominal SAT volume / VAT volume + abdominal SAT volume + total thigh fat-free muscle volume. Abbreviations: body mass index (BMI) body surface area (BSA), intima media thickness (IMT), left ventricular ejection fraction (LVEF), left ventricular end-diastolic volume (LVEDV), left ventricular end-systolic volume (LVESV), left ventricular end-systolic volume indexed to body surface area (LVESVi), left ventricular stroke volume (LVSV), systolic blood pressure (SBP).

| | All-cause mortality | Cardiovascular mortality | Myocardial infarction | Ischaemic stroke |
|---|---|---|---|---|
| **Total population** | | | | |
| *Normal* | 4.62 (4.52-4.72) | 0.70 (0.66-0.74) | 0.88 (0.84-0.93) | 0.48 (0.44-0.51) |
| *Overweight* | 5.47 (5.37-5.57) | 1.03 (0.98-1.07) | 1.44 (1.40-1.50) | 0.67 (0.63-0.70) |
| *Obese* | 7.21 (7.07-7.36) | 1.65 (1.58-1.72) | 1.85 (1.77-1.92) | 0.87 (0.82-0.92) |
| **No Diabetes** | | | | |
| *Normal* | 4.48 (4.38- 4.58) | 0.66 (0.63-0.70) | 0.86 (0.82-0.91) | 0.46 (0.42-0.49) |
| *Overweight* | 5.15 (5.06-5.25) | 0.92 (0.88-0.96) | 1.37 (1.32-1.42) | 0.62 (0.59-0.66) |
| *Obese* | 6.19 (6.05-6.33) | 1.30 (1.23-1.36) | 1.60 (1.53-1.68) | 0.74 (0.69-0.79) |
| **Diabetes** | | | | |
| *Normal* | 14.12 (12.74-15.65) | 2.79 (2.23-3.50) | 2.28 (1.77-2.95) | 1.80 (1.35-2.41) |
| *Overweight* | 13.45 (12.70-14.24) | 3.69 (3.31-4.11) | 3.33 (2.97-3.74) | 1.73 (1.48-2.03) |
| *Obese* | 15.75 (15.10-16.42) | 4.54 (4.21-4.90) | 3.91 (3.59-4.26) | 1.94 (1.72-2.19) |

**Table S4 –** Absolute unadjusted rates of all-cause mortality, cardiovascular mortality, myocardial infarction, and stroke per 1000 person-years of follow-up according to diabetes and ethnicity adjusted BMI category. Data are rates per 1000 person-years (95% CI). Abbreviations: body mass index (BMI); confidence interval (CI).

| | Myocardial infarction | | Ischaemic stroke | |
|---|---|---|---|---|
| | **Unadjusted IRR (95% CI)** | **Adjusted IRR* (95% CI)** | **Unadjusted IRR (95% CI)** | **Adjusted IRR* (95% CI)** |
| **Comparison** | | | | |
| Normal + non-diabetes | 1.00 (reference) | 1.00 (reference) | 1.00 (reference) | 1.00 (reference) |
| Overweight + non-diabetes | 1.58 (1.49-1.69, p<0.001) | 1.27 (1.20-1.36, p<0.001) | 1.59 (1.49-1.69, p<0.001) | 1.15 (1.05-1.26, p=0.002) |
| Obese + non-diabetes | 1.86 (1.73-1.99, p<0.001) | 1.61 (1.51-1.73, p<0.001) | 1.86 (1.73-1.99, p<0.001) | 1.48 (1.34-1.63, p<0.001) |
| Normal + diabetes | 2.64 (2.04-3.44, p<0.001) | 1.61 (1.24-2.09, p<0.001) | 2.65 (2.04-3.44, p<0.001) | 2.49 (1.84-3.35, p<0.001) |
| Overweight + diabetes | 3.87 (3.41-4.39, p<0.001) | 2.09 (1.84-2.37, p<0.001) | 3.87 (3.41-4.39, p<0.001) | 2.23 (1.87-2.66, p<0.001) |
| Obese + diabetes | 4.53 (4.10-5.01, p<0.001) | 2.93 (2.64-3.23, p<0.001) | 4.53 (4.11-5.01, p<0.001) | 2.92 (2.54-3.37, p<0.001) |

**Table S5 –** Unadjusted and adjusted incidence rate ratios (IRR) for myocardial infarction and ischaemic stroke obtained from multivariable Poisson regression analysis in patients grouped by diabetes and ethnicity adjusted BMI category. *Adjusted for age, sex, ethnicity, and smoking status. Abbreviations: body mass index (BMI); confidence interval (CI); incident rate ratio (IRR).

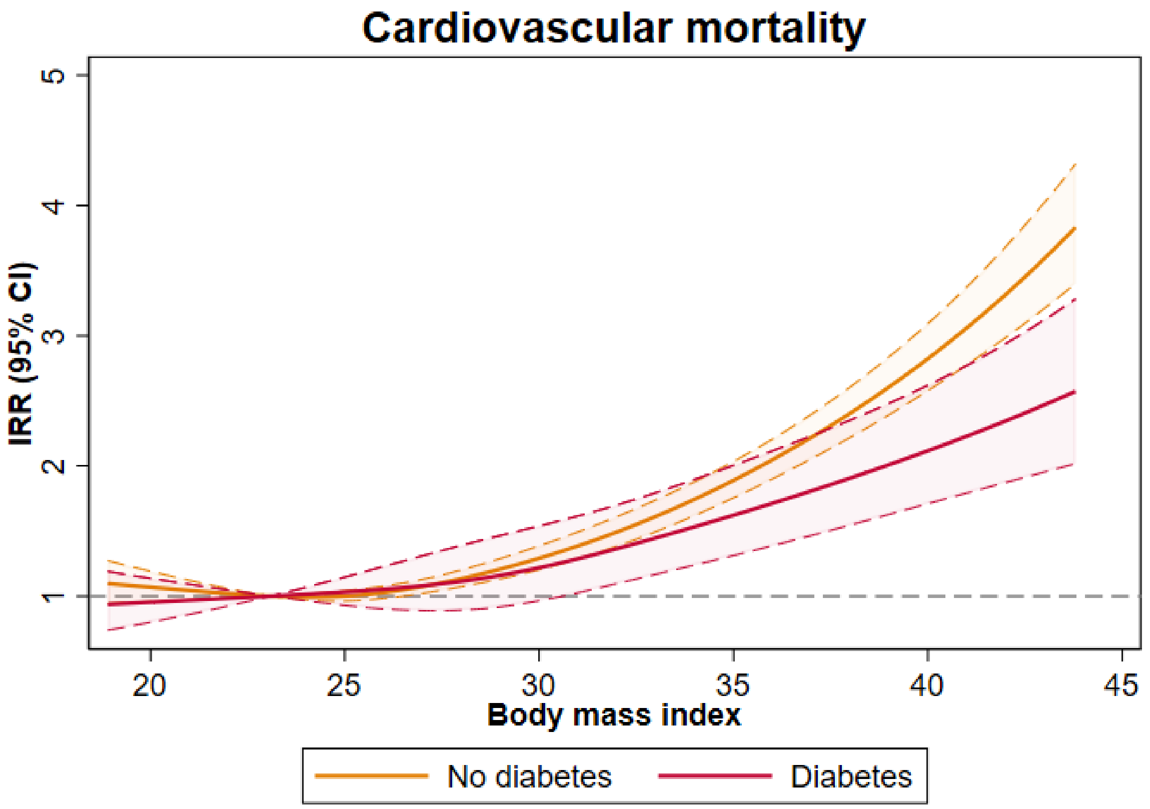


**Figure S1. Cardiovascular mortality according to body mass index and diabetes status.** Incidence rate ratios (IRR) and 95% confidence intervals (95% CI) for cardiovascular mortality according to body mass index (BMI) and diabetes status modelled using restricted cubic spline regression with 4 knots; the reference knot is at BMI 23.4 (median value of cohort). Solid lines represent IRR and shaded areas represent 95% CIs. Spline curves were truncated at the 1st and 99nd centile.
